# Directional Drift in Biologically Meaningful Vector Planes: A Proposed Geometric Framework for Early Detection of Subthreshold Disease

**DOI:** 10.1101/2025.06.05.25329078

**Authors:** Gaurav Prakash

## Abstract

**Background:** Most conventional diagnostic systems rely on fixed thresholds to differentiate disease states from normal. However, early pathological changes may begin before these thresholds are crossed. Therefore, a system that works in this pre-threshold state can potentially lead to earlier diagnosis.

**Objective:** To propose and evaluate a geometric framework that models early disease as a directional drift from a physiological plane to a pathological plane, allowing for pre-threshold detection using biologically interpretable variables.

**Methods:** In this modeling study on synthetic data derived from published clinical trends, two clinically meaningful variables were used to define a 2D feature space. The model was applied to a synthetic dataset of 4000 eyes divided into four phenotypes: normal stable (NS), early disease stable (ED_S), early disease progressive (ED_P), and pre-threshold progressive (PT_P). A physiological plane was constructed using range-normalized values from the NS group. A canonical disease vector was derived from the ED_P group. Each subject’s follow-up data was transformed into a subject-specific drift vector, and the Composite Drift Score (CDS) was calculated as the product of directional alignment (Directional Emphasis Multiplier, DEM) and a Magnitude-to-Noise Ratio (MNR).

**Results:** CDS increased significantly over follow-ups in both ED_P and PT_P, distinguishing them from the two stable cohorts (p<0.001). DEM and MNR components showed consistent trends, with progressive cases exhibiting higher alignment with the disease vector and supra-noise magnitude of change. Visual and statistical analyses confirmed early drift detection even within numerically normal ranges.

**Conclusion:** In this early modeling study based on simulation data, we could quantify the directional drift with a unitless, interpretable metric (CDS) and its derivatives. It showed similar trends in pre-threshold groups as early disease groups, showing potential for further evaluation.

## Introduction

Over the past few decades, there have been significant improvements in diagnostic technology, particularly in imaging and laboratory methods. This has allowed clinicians to detect increasingly subtle physiological variations, even in individuals without overt disease [1–5]. Despite these capabilities, many diagnostic systems continue to depend on fixed thresholds to define disease. This approach can overlook earlier physiological changes that precede those cutoffs [6–14].

### Threshold-based testing

Due to their underlying design, these threshold-based, cut-off style tests tend to perform well in structured datasets where subjects are clearly categorized as either normal or diseased. However, in real-world clinical settings, normal and diseased cases often share overlapping characteristics across multiple parameters. [6,7,13,14]. Oversights with multivariable threshold style testing can lead to redundancy and [15-23].

### Thresholds miss directionality

While threshold-based tests are useful for categorizing disease at a single point in time, they cannot account for how a subject reached that point **(Figure 1 A, B)**. For example, in Figure **1A**, patient P1 remains stable and above the threshold (disease zone) at both *t* and *t* + *Δt*, while P2 worsens but only crosses the threshold at the second time point. Despite their very different disease dynamics (P1 has stable disease and P2 has worsening disease), both appear similar if only the current value is considered. Also, deviation away from normal state and progression of worsening often begins well before any statistical cutoffs are breached. For example, **Figure 1B** shows two more patients, P3 and P4, coming for their first-ever visit at time t. Both are within the so-called normal range currently. However, P3 has been drifting steadily toward the disease state from before, while P4 is stable. A static threshold cannot distinguish between them, failing to detect the early directional trend in P3. Early pathological drift begins when the subject or the organ is still numerically within the accepted values for ‘normal’ population distribution. This phenomenon has been documented in many chronic progressive diseases [24-28].

**Figure 1:**
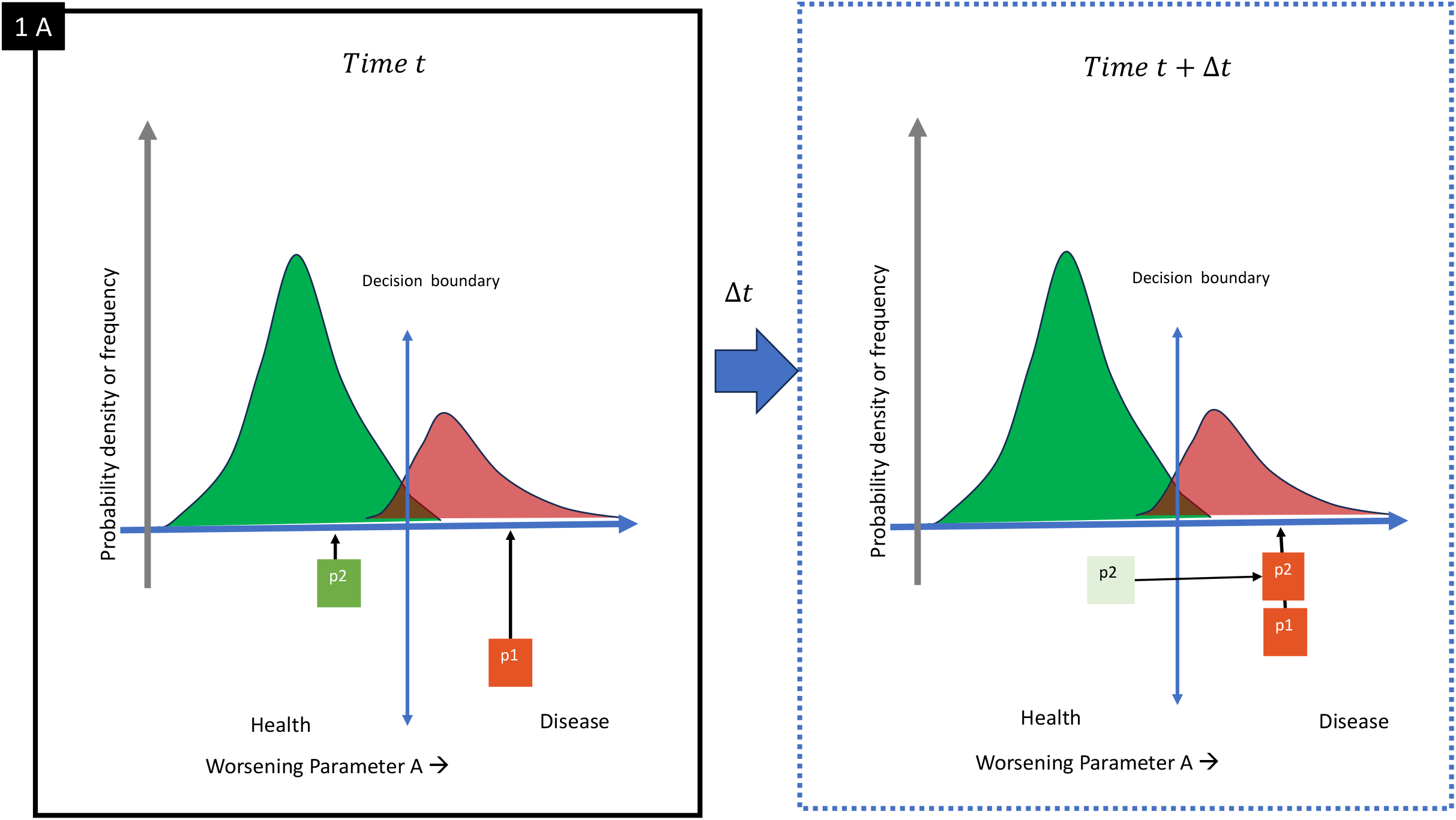
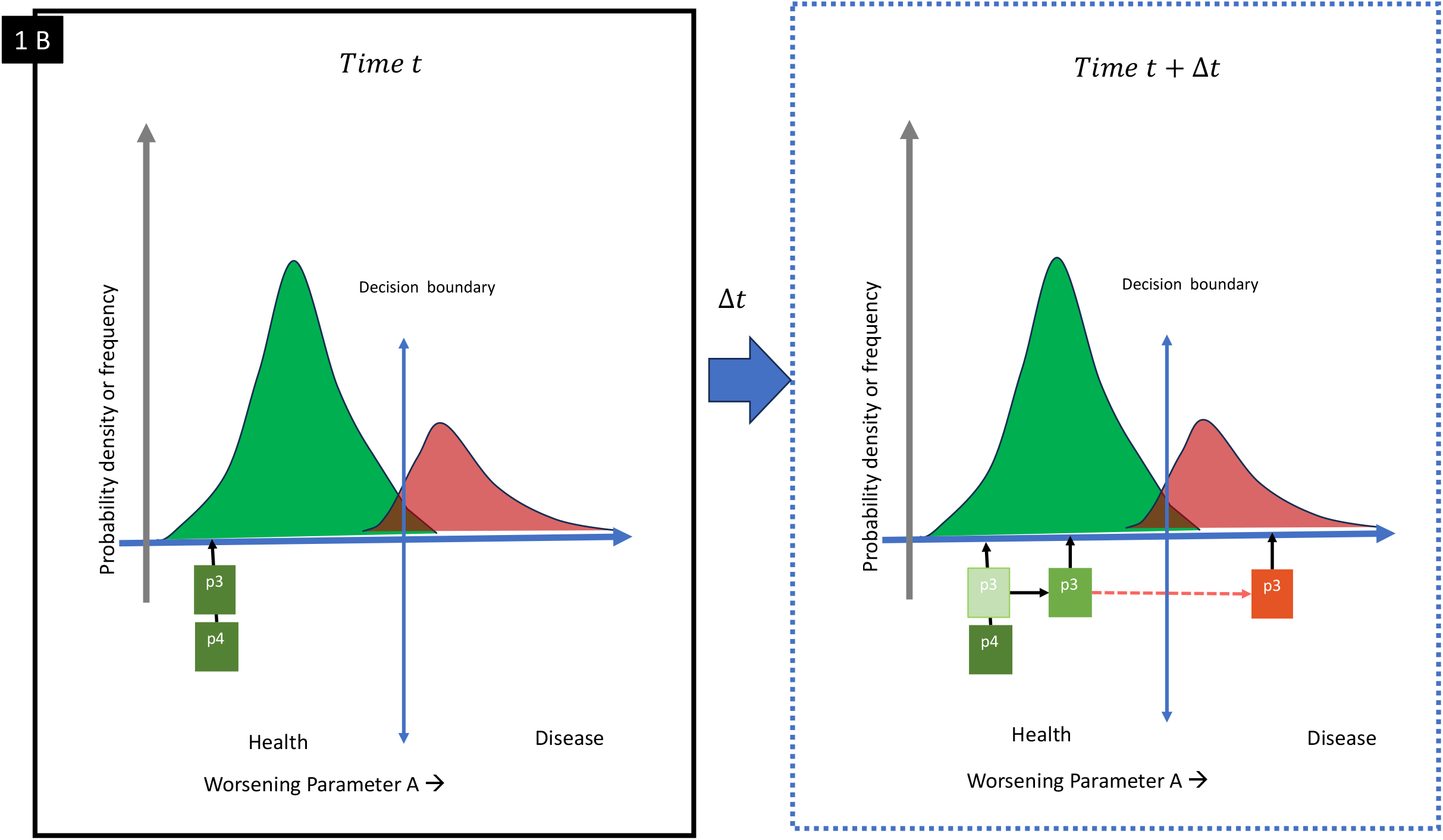
The trend missed in threshold-based tests. **Figure 1A**. Two patients in the disease zone may appear clinically similar at a single timepoint despite very different trajectories leading up to disease. P1 is stable and P2 is worsening. **Figure 1B**. Two subjects in the normal zone may be on very different paths. P3 is stable but P4 was worsening.

Clinicians have long recognized that normal range, sub-threshold rising values (more than the reference change value), can be concerning [29-31]. However, it has been challenging to formally characterize or quantify this type of change using traditional tools. Worsening this issue is the assumption that physiological parameters follow Gaussian distributions, which is often not accurate [7,32-34]. Cross-sectional cutoffs are further complicated by changes in how these scores, or numerical thresholds are defined: as clinical panels revise cutoffs, the decision boundary itself moves one way or the other [30,35]. One such possible concept is to detect drift: a directional deviation away from the normal state. To explore the potential of using drift as an alternate vantage point, we should first review natural fluctuations (to due homeostasis) and observation-related noise and formalize our definitions with reference to this study.

### Homeostasis, physiological variation, observation variation, noise, and drift

#### Homeostasis

Healthy physiological systems maintain a stable state through active regulation resulting in natural fluctuations. Homeostasis is the organism’s tendency to maintain this state.

#### Physiological variation

The homeostatic self-correction oscillations occur within bounded ranges. [36-38].

*Observation variation*, on the other hand, arises due to multiple reasons within the device-operator-interpreter system. This occurs whenever a measurement is attempted on a biological system or structure [39-46]. *Observation variation*, like *physiological variation*, is typically non-directional and self-correcting.

#### Noise

Physiological and observational variation can interact stochastically. There may be not a clear rule to discriminate them from each other. Therefore, it may be mathematically useful to club them into a single entity, ‘noise’.

*Drift* refers to a slow, directional deviation away from this physiological corridor aligned with the expected direction of pathology progression. This occurs before conventional diagnostic thresholds are crossed. It is not defined by a single measurement but by a trajectory over time. This is often still within the so-called “normal” numerical range. [47-55].

It can be argued that medicine, in essence, is systems biology viewed through the lens of pathology. In systems biology, it is well recognized that transitions in system behavior are often preceded by early warning signals. These signs include increased variance, autocorrelation, or critical slowing down, where recovery from oscillations becomes delayed [56-59]. These signals suggest that a system is approaching a tipping point, after which a spontaneous return to the original state becomes unlikely.

Therefore, evidence, as outlined above, suggests homeostasis and drift state are not similar, following different trajectories and presentations. These two states therefore appear to be governed by different sets of rules which can be potentially utilized to identify them. The simulated temporal and trajectory-based behavior of 6 representative cases (including one ‘stable normal’) illustrates this further in Figure **2 A-D**. In this visualization, we show six representative simulated cases: one stable normal and five with varying disease trajectories **(Figure 2A)**. These illustrate the diverse ways patients can drift from physiological equilibrium. Some progress gradually, some abruptly, and some not at all. Highlighting the contrast between normal noise (green zone) and true disease drift (blue curve) can suggest how directional movement outside the noise corridor marks meaningful change even before the threshold breach **(Figure 2B)**. Cross-sectional views can obscure these temporal patterns. **Figure 2 C** demonstrates this by comparing overlaid distribution plots at t_0_ to t_5_. Despite clear progression in several cases, static thresholds at single time points miss the bigger picture. This creates a zone of clinical opportunity zone: a region of directional drift where early intervention could alter trajectory (**Figure 2D)**. Case 5 in particular shows how an early flag could precede and prevent threshold-defined diagnosis. This case becomes stable after early pre-convectional threshold diagnosis and treatment.

**Figure 2.**
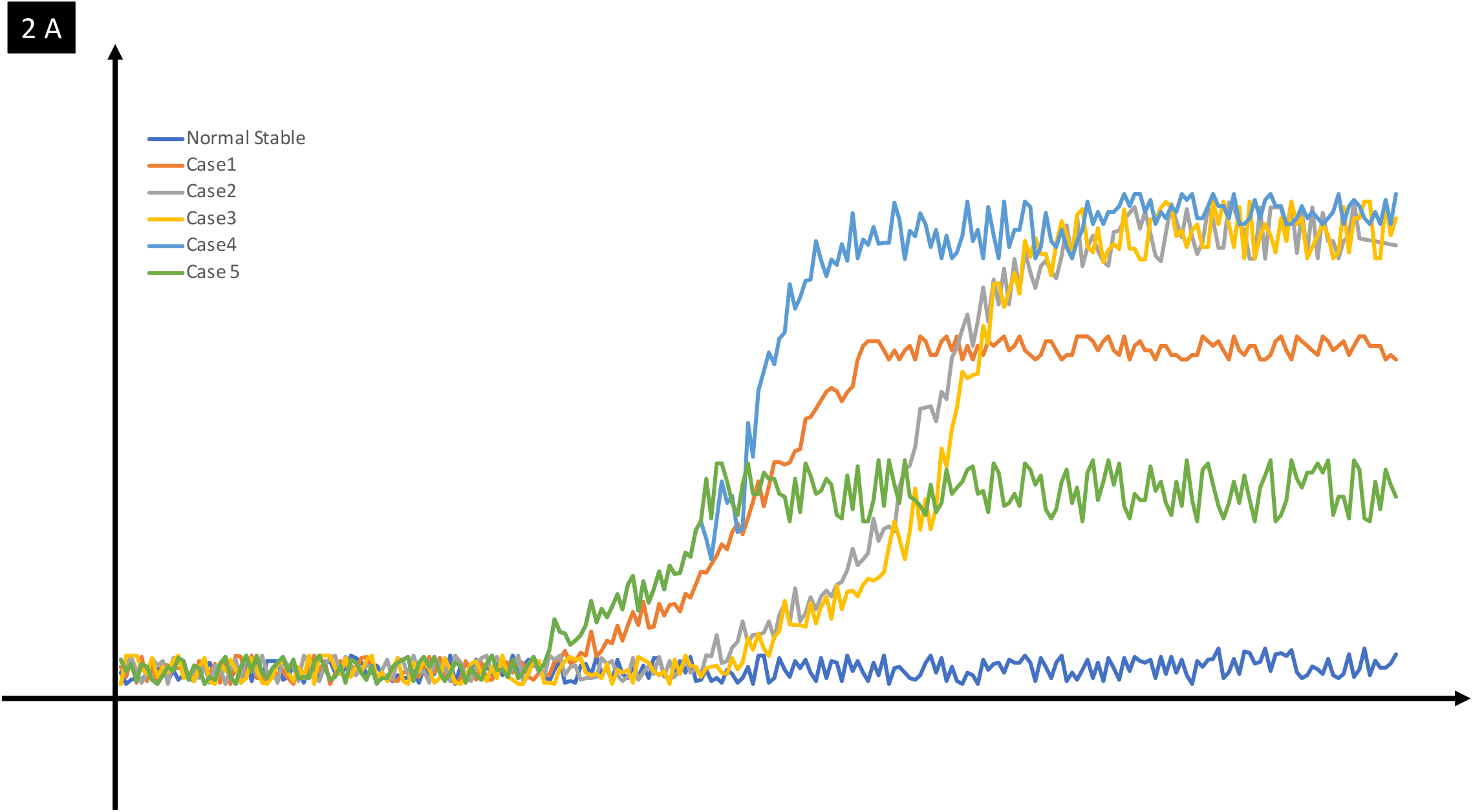
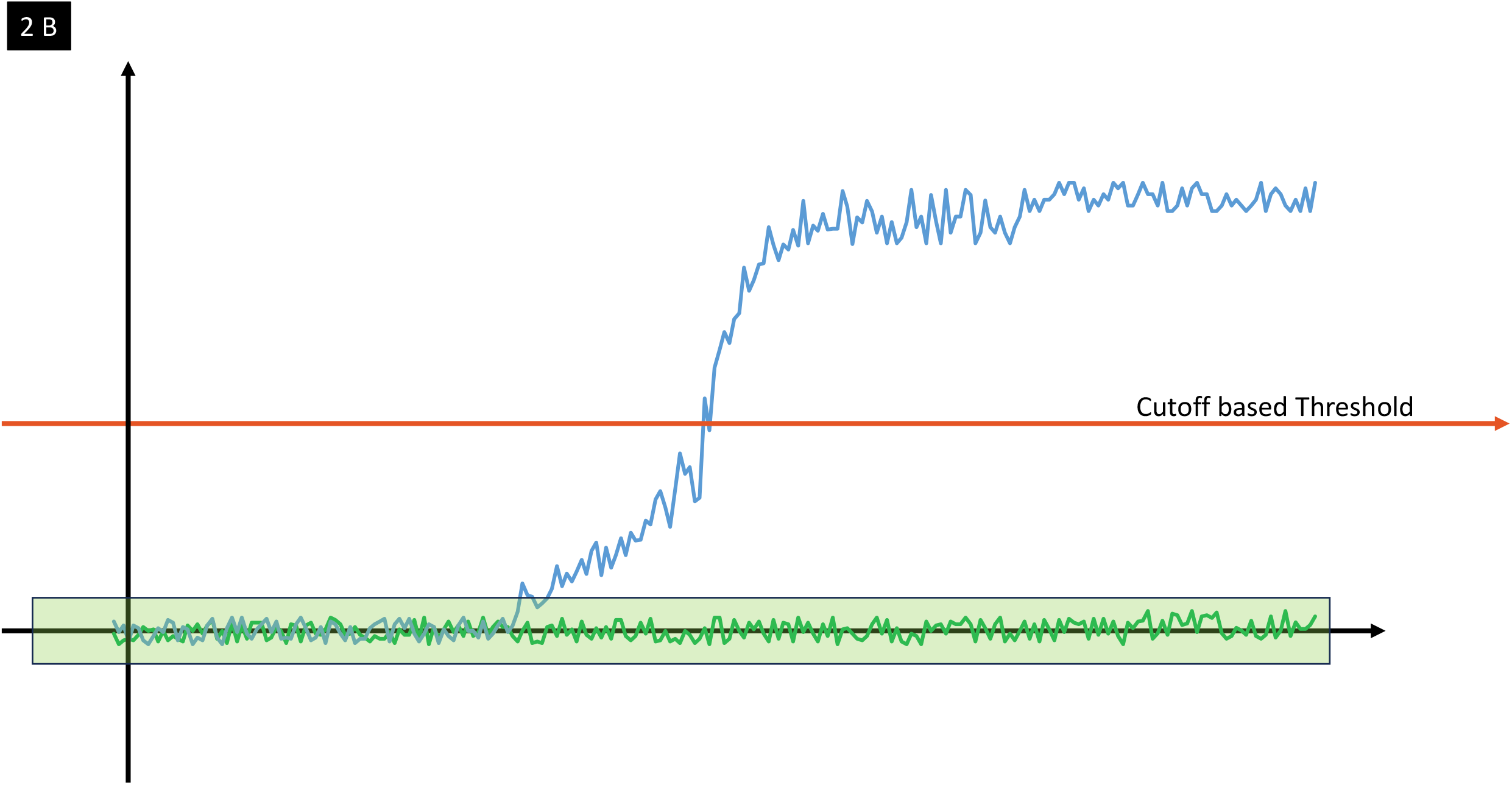
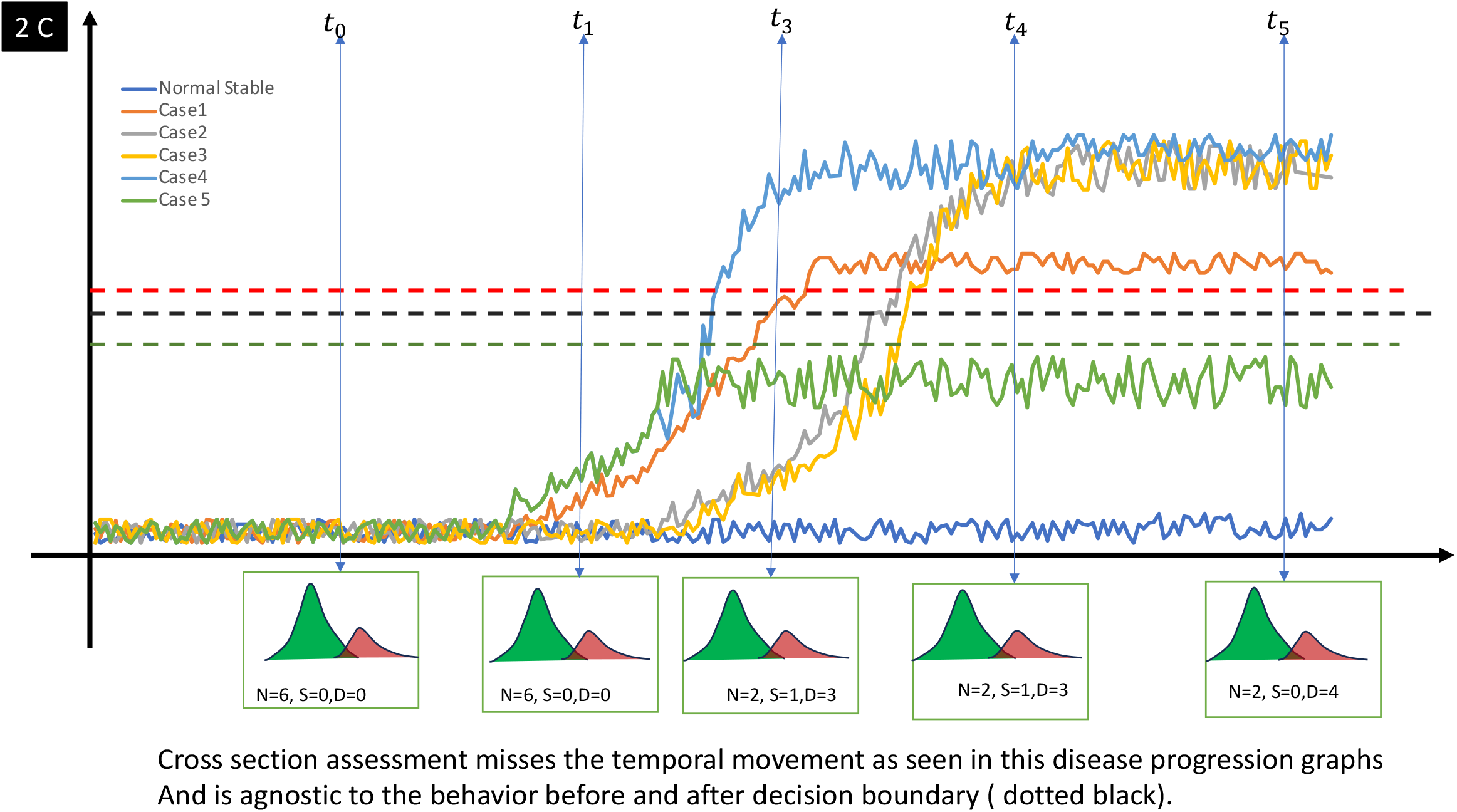
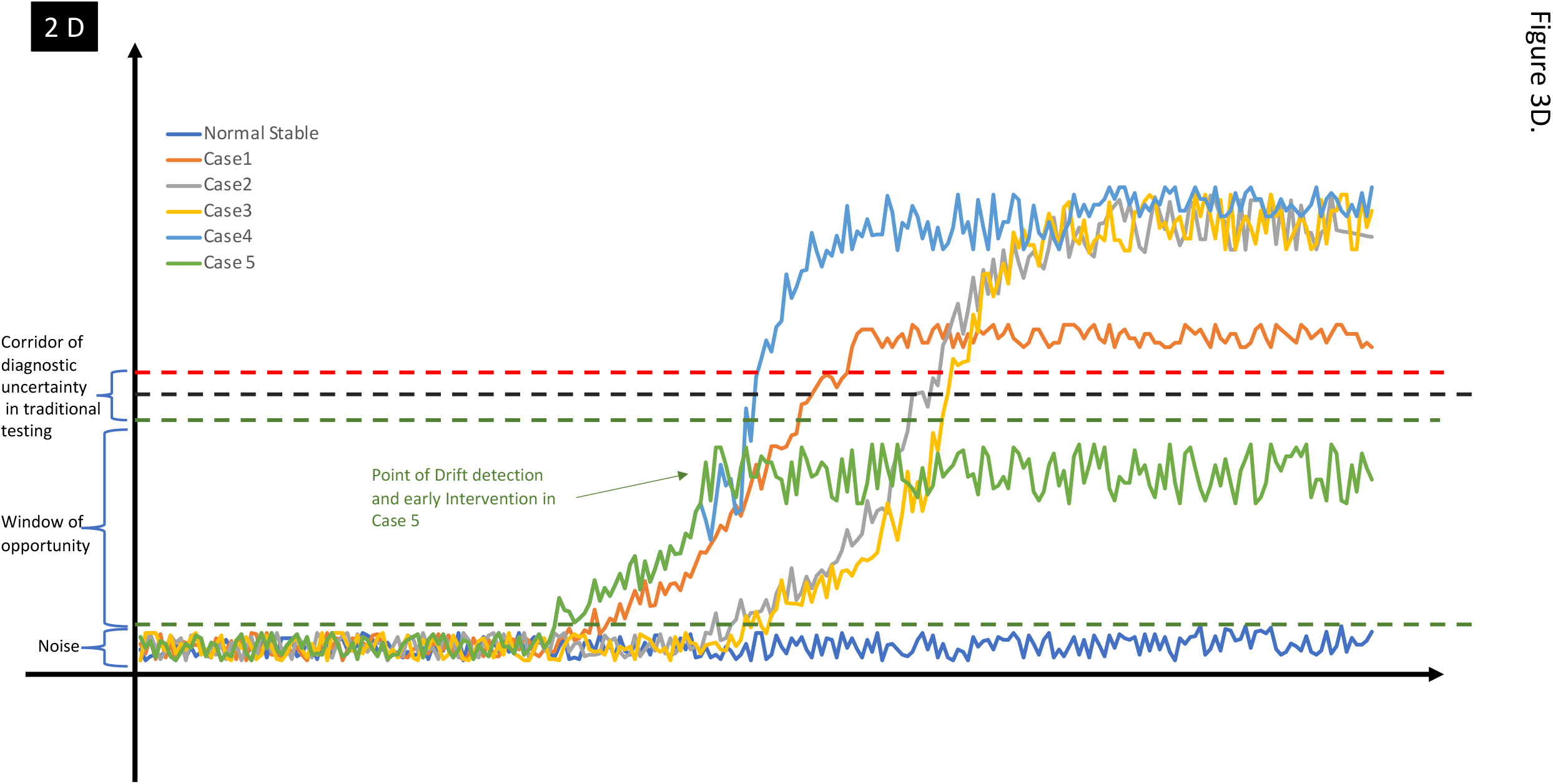
A–D. Temporal patterns of directional drift in simulated subjects. **Figure 2A**. Six representative cases showing variable progression and stability. **Figure 2B**. Comparison of noise corridor vs disease trajectory. **Figure 2C**. Timepoint-overlaid cross-sectional views demonstrating missed progression (in the distribution plots, N= normal, S= suspect, D= Disease) **Figure 2D**. Conceptual “window of opportunity” for early detection and intervention.

#### Mapping the trajectory on a plane and not a cross-sectional cutoff

As we have discussed earlier, traditional diagnostic frameworks assume that normal and disease lie along a shared linear axis. This implies that progression is merely a matter of movement along a single continuum [6,7,60,61]. It is a mathematical requirement for comparison and is therefore convenient [60-62]. Threshold-based models assume that a variable’s behavior is interpretable within a known distribution. However, when variables come from fundamentally different biological states, such as a regulated homeostatic system compared to a decompensating one such as the drift, this assumption becomes sub-optimal.

#### Two plane hypotheses

We propose that normal and disease states are better represented as existing on distinct geometric planes. The physiological plane is constrained by homeostasis. Values fluctuate but remain bounded and self-correcting. The disease plane has different behavior, what we term as ‘intentional directionality’ to differentiate from noise (which on measurement shows as non-intentional, non-directional movement). The tipping point marks the origin of the disease plane **(Figure 3)**. This transition is illustrated in the Figure 3, where normal subjects remain within the physiological plane with the passage of time, while progressively worsening pre-threshold and diagnosed cases drift on the pathological plane after the tipping event. The overlay of threshold-based distribution panel on the left of the image illustrates how classical models may produce overlapping decision regions, whereas the geometric model separates populations through angular divergence between planes. The exact moment of tipping may not be directly observable when intermittent sampling is being done compared to real-time tracking. In a clinical setting, this means that the tipping point may occur between follow-ups. However, the behavior before and after the tipping point can be traced back to a putative inflection point, giving mathematical support to this concept. Once tipping occurs, the subject may still appear within the normal range numerically. However, its new trajectory begins to follow the rules of the disease plane **(Figure 3)**. The parameters continue to change and eventually cross conventional diagnostic thresholds, progressing further into disease.

**Figure 3:**
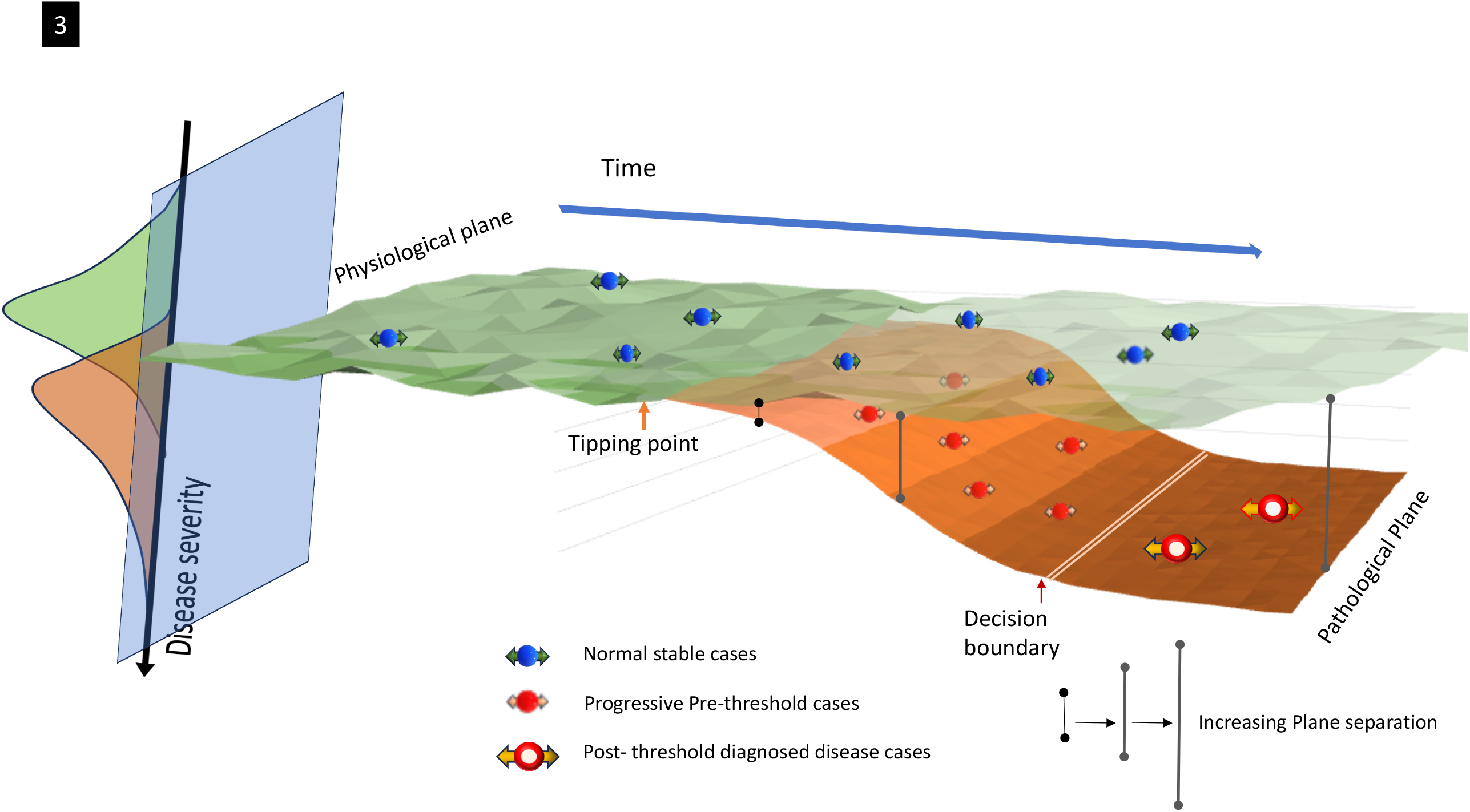
The two plane hypotheses and directional drift concept. Conceptual model of the two-plane hypothesis. Normal subjects remain within the physiological plane, while progressive cases drift onto the pathological plane after the tipping point.

#### Benefits from formal quantification of this concept

The concept of the two planes can be extended beyond qualitative or illustrative value by mathematically defining them. This can help in creating a framework that documents the changes occurring on these planes around the tipping point. This way, it can be attempted to extend to clinical implications. In the next section, we present a vector-based geometric framework that captures this transition in feature space.

## Methods

For this exploratory study, we first developed a geometric framework in which normal and disease planes, and their derived vector products, are defined. We then formalized the rules governing their interactions and applied this framework to a simulated dataset, built on biological behavior described in literature:

A. **Framework design:**
  1. Variable selection and feature space definition: Let *x* and *y* denote two biologically meaningful variables reflecting the disease process, which meet the following criteria:
    a. Established clinical relevance in the disease process.
    b. A biologically plausible explanation for their behavior in the disease progression with the presence of directionality
    c. Widely available and commonly used in clinical practice.
    d. Whenever possible, selected variables should be simple, direct measurements or low-complexity derived values with a clear biological rationale.
    e. Low correlation in health subjects
  2. Normalization for physiological (normal) subjects: Once identified, the values for these variables are first extracted from a dataset representative of physiological (normal) cases. The two variables are then range normalized:
    a. Physiological Range was calculated as

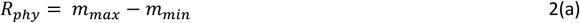 Where *m* ∈ {*x, y*}, the original(raw) value of the variable from physiological cases.
    b. If the variable’s value is known to increase with disease progression

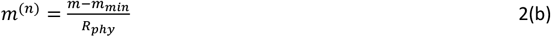
    c. If the variable’s value is known to decrease with disease progression

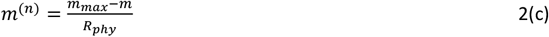 Thus, normalization is performed according to the transformation rules above resulting in the directionally aligned normalized variables *x*^(*n*)^ and *y*^(*n*)^ Where: *m* ∈ {*x, y*}, and,
      - *m* ∈ {*x, y*} is the original value of the variable in the physiological cases.
      - *m*^(*n*)^ is the normalized value for physiological(normal) cases.
      - *m*_*max*_, *m*_*min*_ are the maximum and minimum values seen in the physiological population. *Note: Superscripts in parenthesis in this manuscript denote indices (e*.*g*., *normalized) and should not be interpreted as exponentiation*.
  3. Creation of Physiological plane: This creates a bounded plane defined by normalized coordinates with multiple physiological subjects occupying positions in this coordinate system.

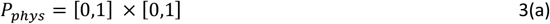

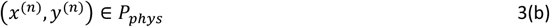 Where:
    - *P*_*phys*_ is the physiological (homeostatic) plane.
    - *x*^(*n*)^, *y*^(*n*)^ are the normalized values of the two selected variables, each scaled to lie within [0,1].
  4. Scaling for pathological (diseased) subjects: The same two variables *x* and *y* are extracted from the dataset of patients with early to moderate disease. We exclude patients defined as borderline or suspect to map established disease behavior. Each variable is then scaled by the range noted in the physiological set, using the following transformation rules:
    a. If the variable’s value is known to increase with disease progression:

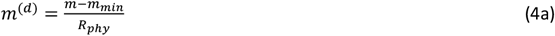
    b. If the variable’s value is known to decrease with disease progression:

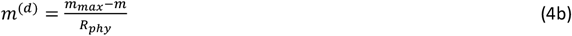 Thus, *scaling (not normalization)* is performed according to the transformation above resulting in the directionally aligned scaled variables *x*^(*d*)^ and *y*^(*d*)^ Where: *m* ∈ {*x, y*} and,
      - *m* ∈ {*x, y*} is the original value of the variable in the pathological cases.
      - *m*^(*d*)^ is the *scaled* value.
      - *m*_*max*_, *m*_*min*_ are the maximum and minimum values seen in the <u>physiological</u> population as described in the normalization section.
      - *R*_*phy*_ is the physiological range, as defined in Equation 2(a)
  5. Creation of the pathological plane: in contrast to the physiological plane, this scaled plane extends in the direction of disease progression and is eventually bounded by the End state (*ε*, the most severe disease state biologically observed before complete loss of structure or function).

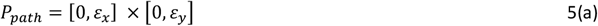

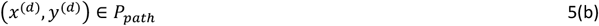 Where:
    - *P*_*path*_ is the pathological (post-tipping event) plane.
    - *ε*_*x*_, *ε*_*y*_ are the scaled disease state maxima derived from the known extreme of the disease, or the End state.
    - *x*^(*d*)^, *y*^(*d*)^ are the scaled values of the two selected variables. The scaled disease state maxima (*ε*_*x*_, *ε*_*b*_) serves only to define the upper edge of the pathological plane; since this study focuses on early drift just beyond the physiological zone, we do not model scenarios where *x*^(*d*)^ → *ε*_*x*_ *or, y*^(*d*)^ → *ε*_*y*_ . Pathological subjects are conceptualized to have transitioned to this plane once the tipping point is crossed and are now on this plane. They can occupy any location in this feature space and have inter-measurement variability much like the physiological subjects. More importantly, they are expected to move in this coordinate plane towards the direction of worsening pathology unless they have achieved stable disease status showing what we define next as ‘*meaningful directionality’*.
  6. Quantifying ‘*meaningful directionality’*: The first step is identifying *<u>consistent</u>* deviation beyond expected oscillation in a subject initially labeled normal. When this deviation is aligned with the direction of disease progression, it can be interpreted as movement characteristic of the pathological plane (or ‘*meaningful directionality’*). This suggests early disease transition in that subject independent of its distance from the classical, cross-sectional cutoff based, decision boundary. The next step is to quantify this meaningful directionality and create a framework for testing unlabeled cases. For this, we define the following:
    - A pooled noise scalar: ***η***
    - A pooled disease vector: 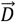
    - A subject-specific drift vector: 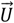
      a. Noise: The noise is a scalar ***η*** modeled from the expected physiological noise derived from pooled within-subject standard deviation (*Sw*) over the visits for physiological patients. For this study, one measurement per visit per patient was modeled.
        i. For each physiological subject *j* ∈ {1,2, …, *J*} the within-subject standard deviation is calculated using all visits *i* ∈ {1,2, … *q*_*j*_}, where *q*_*j*_ denotes the number of follow-up time points available for subject *j* . The standard deviation for each normalized variable *x*^(*n*)^, *y*^(*n*)^ is calculated as:

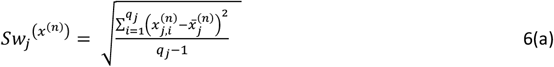

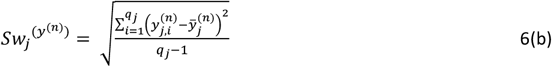

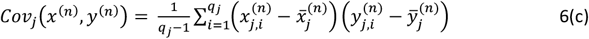 Where:
          - 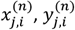 represent the *i* − *th* visit for subject j.
          - 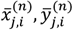 are the subject-specific means.
        ii. These values were used to compute the joint within-subject standard deviation for every physiological subject *j* as:

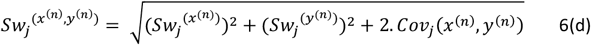
          1. Final Noise Magnitude: The coefficient of repeatability (*CR*), representing the 95% confidence interval for repeated measures for a single variable, is calculated as [63]:

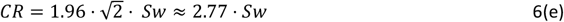 In our framework, we extend this concept to the multivariate case, where *Sw* values are pooled across the two axes. The final scalar noise magnitude is therefore defined as:

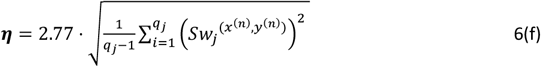 This constructs a radial 95% confidence boundary around expected physiological drift in this joint feature space. While this is conceptually an extension of *CR* used often to define the agreement between devices, its operational role here differs. It serves as a signal-to-noise denominator in a vector framework thereby providing clinical threshold guidance. This is an extension of its previous application in a univariate case as we used in a previous study [64].
      b. Age-related, physiological reference drift: This maps the directionality of the true labeled normal (physiological) cases. Many physiological parameters change with time due to natural aging response and this needs to be mapped to differentiate from pathological change. This can be affected by multiple processes including the age of the subject [65-67]. However, the changes over a shorter period such as less than a decade (compared to entire lifetimes) can be modeled using a linear approximation, especially for exploratory study. This rate of change has been described in multiple longitudinal studies. The resultant vector is:

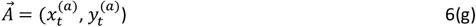 Where:

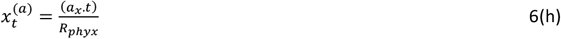

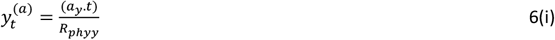 With:

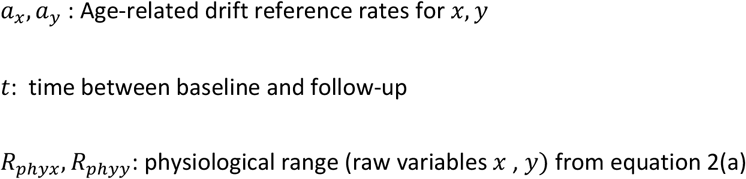 For this exploratory study, we assumed the drift-related change over the 2.5 years we sampled to be negligible. However, the equations were constructed including this drift to keep a placeholder for future iterations of the model.
      c. Disease vector: This maps the directionality of the true labeled disease (pathological) cases compared to the physiologically expected (reference) drift. Disease progression can be non-linear over the entire spectrum of pathology. However, the current model is meant for early detection in a population hitherto labeled as normal. The closest approximation for that rate of change seems to be cases with early but established disease (or lower-grade disease) showing progression.
        i. For each pathological subject *k* ∈ {1,2, …, *K*}, the scaled change between baseline 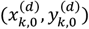 and follow-up visit 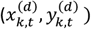, for all *t* ∈ *T*_*k*_ is calculated adjusting for age-related physiological reference drift 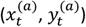. Here*T*_*k*_ denotes the set of valid follow-up time points for subject *k*, and *K* is the total number of pathological subjects included in the disease vector calculation.

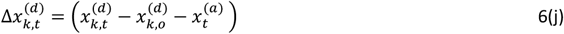

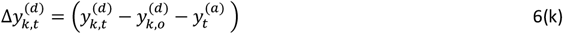
          2. Per-subject mean disease trajectory vector: For pathological subject *k*, we compute the average of these vectors across all their follow-up visits:

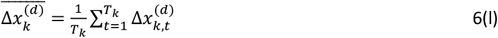

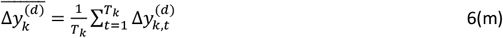
        ii. These subject-level vectors are then averaged to define the **pooled disease vector**.

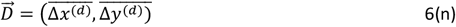 Where:

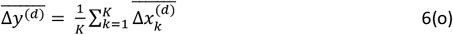

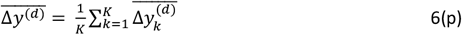
        iii. the angle of the pooled disease vector 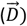 in reference to the *x, y* Cartesian frame is:

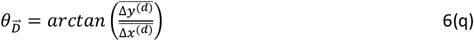 The resulting disease vector 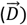 thus serves as the reference direction against which subject-level drift vectors are compared.
      d. **Subject drift vector:** For any subject under evaluation (denoted as unknown subject *u*) at follow-up visit *t* after a baseline visit *t*0 to be evaluated and compared to the pooled metrics of noise and disease vector as above, the following method is performed:
        i. Scaling done on raw *variables* (*x*_*t*_, *y*_*t*_,) to create scaled variables for both time 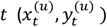 and baseline *t*0 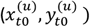 using transformation principles form Equation 4(a) and 4(b).
        ii. The change between scaled values at baseline 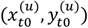 and follow-up 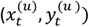 at time *t* can be calculated, adjusting for physiological reference drift 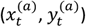 as:

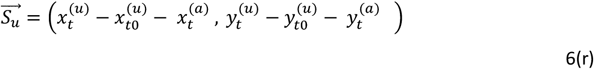
        ii. The magnitude of the subject vector is:

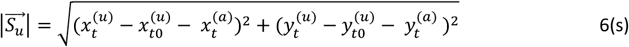
        iii. The **angle** 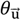 of the subject vector is:

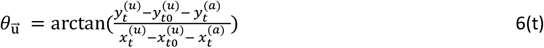
  7. **Metrics to quantify meaningful directionality and establish a decision threshold:** In the sections above, we constructed the framework of two reference planes: a physiological plane, *P*_*phys*_ = [0,1] × [0,1]), and a pathological plane, *P*_*path*_ = [0, *ε*_*x*_] × [0, *ε*_*y*_] (see Sections 3a and 5b). We also defined a noise metric ***η*** from the data in *P*_*phys*_ (Eq.6f), a canonical disease vector 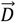 from data in *P*_*path*_ (Eq.6n), a subject-specific drift vector 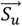 (Eq. 6s) based on scaling principles from *P*_*path*_. We now introduce three derived metrics to evaluate a subject’s deviation from normal behavior (in magnitude) and alignment toward disease-like behavior:
    i. **Change greater than physiological noise:** If a subject has drifted more than that expected by normal physiological oscillation, that becomes the first indicator for concern. Traditionally, limits of repeatability have been applied in a subtractive form (e.g., change > 2 Sw), but such thresholds depend on variable’s units (e.g., D (diopters), *μ*m, mg/dl, etc.) and reduce cross variable or cross-domain generalizability. Ratios, on the other hand, are unit agnostic and therefore intuitive to compare. Therefore, we next define the Magnitude to Noise Ratio:

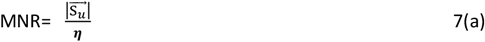 In its essence, this is a signal-to-noise (SNR) ratio and is an operational metric. Just as a radio-based system uses SNR to deduce a meaningful signal over background static, the MNR intends to deduce subject-level change over the expected noise.
    ii. **Directional alignment to disease process:** for the unknown subject *u* being evaluated, the cosine of the angle between the subject’s drift vector 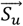 and the disease vector 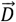 is computed as

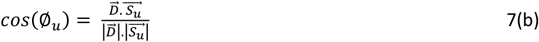 This value captures the angular alignment between the subject’s direction of change and the pooled disease trajectory. For this comparison, a perfectly aligned subject would have *cos*(Ø_*u*_) = 1, while a subject drifting orthogonally or in the opposite direction would have values approaching 0 or negative. Medicine works significantly on pattern recognition and disease phenotypes tend to get replicated in areas of feature space that are proximal to each other. Therefore, we further applied a signed directional weighting. We call this **Directional Emphasis Multiplier**

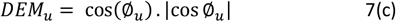 In this study, we used a signed square of the cosine between the subject and disease vector directions. The goal is to produce a curve that favors well-aligned movement and suppresses non-aligned vectors. However, since this metric is customizable, it can be modified based on real-life variations.
    iii. The final score combines both directional and magnitude components: The **Composite Drift score** is:

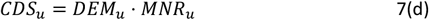 This scalar, unitless value reflects both the strength and directionality of pathological drift and forms the basis for early detection in this framework. It is intuitively set at a threshold of ≥1.0 (perfect alignment with disease process and change greater than seen with noise, which can vary over multiple case scenarios as we discuss later in results and discussion).
  8. **Expansion to multidimensional space and adjustments for high covariance:** For a ≥3-dimensional model, or in cases where there is a significant covariance, the above method may be challenging even after scaling. We explored the candidate methods for such situations. We evaluated Mahalanobis distance (MD) as a prospective alternative method to set up this for future iterations of this model [68-70]. CR is used more commonly in repeatability studies and therefore it is more intuitive for clinicians. Mahalanobis distance is used as a robust compensation for the covariance in multiple fields including medicine The two parameters, we used in this study to represent *x and y*, Kmax and TCT do not have significant covariance in normal cases. However, we suspect that this can be a potential concern in some clinical situations, especially when this framework is expanded. Ideally, matrix manipulation is easier in a programming environment such as Python, however, in this exploratory study we wanted to ensure the generalizability and have used Excel environment. In next steps, we outline the method we used to calculate Mahalanobis distance:
    i. For each normal subject j ∈ {1,2, …, *J*}, and for each follow-up point *t* ∈ *T*_*j*_ we used the normalized values obtained from Equations 2a,2b and computed the change relative to baseline for variables *x* and *y* respectively. These were arranged in row vectors:

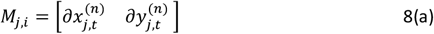 Where: 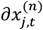 is the change in normalized *x* compared to baseline for subject *j* at follow-up visit *t* 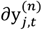 is the change in normalized *y* compared to the baseline for subject *j* at the follow-up visit *t* *(It should be emphasized that this method does not use the baseline visit as a separate visit like when calculating Sw but the difference of baseline from follow-ups)*.
    ii. Aggregating across all subjects and all follow-up time points yielded the following data matrix with all the data points.

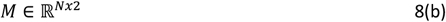 Where:

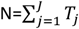
    iii. From this matrix *M* ∈ ℝ^*Nx*2^ we calculated the sample covariance matrix:

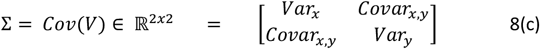 This matrix contains variance of *x* and *y* along the diagonal and covariance between *x* and *y* off-diagonal.
    iv. For each subject under evaluation (denoted as unknown subject *u*) at follow-up visit *t*, we define their normalized drift vector relative to baseline as:

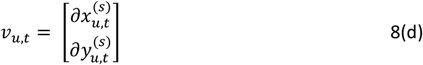 Where: 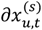 is the change in scaled *x* compared to baseline for subject *u* at follow-up visit *t* 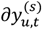 is the change in scaled *x* compared to baseline for subject *u* at follow-up visit *t* (scaling done as in equation 4a,4b, as the status (disease or normal) is unknown)
    v. The Mahalanobis distance (MD) for the change seen in subject *u* at follow-up *t* from the pool of expected change in normal subjects is therefore calculated as:

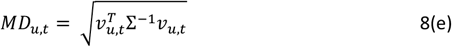 Where: 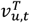 is the transpose of the matrix *v*_*u,t*_ for subject vector from equation 8(d) Σ^−1^ is the inverse of the covariance matrix (Σ)of pooled normal subjects from equation 8(c) The next step is to compare this distance to a reference point. As a guideline, most studies including ours have used 2.77 times *Sw* as *CR* (see equation 6e), which translates into 95% of the observed change within expected variation in a univariate model. However, Mahalanobis distance follows a chi-square distribution. Therefore, we utilize the chi-square distribution for 2 degrees of freedom to derive an equivalent multivariate confidence boundary. The 95th and 99^th^ percentile of this distribution corresponds to approximately 5.99 and 9.21. The square root of these values gives thresholds as below:

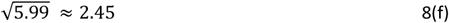

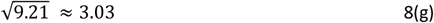
    vi. To compare with our Euclidean distance-based CR model (from equation 6f) we used the scaling logic derived above. We used 2.45 for the initial iteration as this corresponds approximately to the 95% percentile logic used for the CR. This will change based on the degrees of freedom and required alpha and is customizable for future iterations. To keep intuitive comparability with the MNR derived above in 7a, we call this entity.

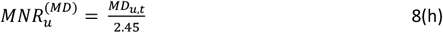
    vii. The CDS hence calculated based on this 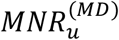 is:

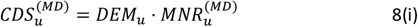 Where:

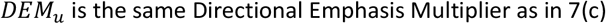
B. **Application to synthetic subjects:** With the above framework established, we applied the above steps 1-8 on a labeled synthetic dataset composed of four distinct subject groups (n=1000 per group), representing a different disease stage or behavioral phenotype. For this exploratory study, we wanted to explore a disease with domain knowledge before expanding to others in future studies, so that we are aware of the expected physiological and pathological behaviors. Therefore, we modeled normal cornea and early keratoconus for this study and the two variables used were steepest Keratometry (Kmax in diopters) and thinnest corneal thickness (TCT) in microns. The synthetic data was generated using normal and abnormal ranges, rate of progression, and intra-measurement standard deviation provided in the literature [71,72]. The exact data generation methodology, including the randomization logic and Gaussian noise parameters, can be shared on request. To prevent any future chances of data redundancy or overfitting, we did not use any of our previous studies or our published data with normative corneal or keratoconus data.
  1. **The 4 subject groups were**
    a. *Stable normal cases:* Used to define the reference range for physiological noise *η* and physiological reference drift 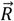.
    b. *Stable (early) disease cases*: served as anchor points for comparison with progressive disease. These cases belong clearly to the pathological plane (no suspects or borderline case) without directional progression.
    c. *Progressive (early) disease cases*: These subjects also belonged to the pathological plane and have with similar baselines as (b). They were simulated to reflect realistic but stochastic disease progression. Changes in the individual variables *x, y* were adjusted to trend over worsening values consistent with known early disease behavior over time. The disease vector 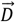 was calculated from this group.
    d. Progressive normal-range cases: This group represents the primary target of our framework: subjects whose baseline values fell within traditional “normal” ranges (based on Gaussian cutoffs or consensus thresholds), but whose homeostatic compensatory mechanisms have failed, leading them to drift beyond the biological tipping point and thus, by definition, into the pathological plane. Variables *x, y* were simulated to reflect stochastic but directionally worsening trends, consistent with known early disease behavior over time. To maintain biological plausibility, the average rate of change in this group was scaled to approximately 80% of that used for the early progressive disease group (Group C). This conservative modeling assumption reflects the current lack of direct empirical data on progression dynamics in this pre-diagnostic window. The drift parameters used here are customizable and can be refined in future models using more granular longitudinal datasets.
  2. Computations in the synthetic dataset: noise (η) and disease vector 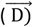 were computed at the dataset level, and 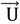,cos(ϕ), DEM, MNR, and CDS were computed at the subject level.

## Results

### A. Group Characteristics and Internal Validation

The synthetic exploratory dataset included four simulated groups (n = 1000 each): Normal Stable (Norm), Early Disease Stable (ED_S), Early Disease Progressive (ED_P), and Pre-threshold Progressive (PT_P).

As noted previously, in this study, the two variables *x and y* were assigned values from Kmax and TCT, respectively, such that Kmax → *x* and TCT → *y*. All subsequent analyses refer to these generalized variables *x and y* for clarity and broader applicability.

Group means and standard deviations for baseline (*t*0) and final follow-up (*t*5) values of (*x, y*) and normalized variables *x*^(*n*)^, *y*^(*n*)^ confirmed expected characteristics hypothesized while modeling the framework. Both the groups designed to worsen with time (ED_P, PT_P) showed worsening of variables with time, while the groups designed to remain stable (Norm, ED_S) remained relatively unchanged **(Tables 1 and 2)**.

**Table 1.** Values (mean ± SD) of raw variables *x and y* at each timepoint (t0 to t5) for all four cohorts.

|  |  | Early Disease<br>Progressive |  | Early Disease<br>Stable |  | Normal Stable |  | Pre_Threshold<br>Progressive |  | Total |  |
| --- | --- | --- | --- | --- | --- | --- | --- | --- | --- | --- | --- |
| Raw<br>variable |  | Mean | Std.<br>Deviation | Mean | Std.<br>Deviation | Mean | Std.<br>Deviation | Mean | Std.<br>Deviation | Mean | Std.<br>Deviation |
| x | t0 | 48.72 | 2.39 | 48.75 | 2.37 | 43.70 | 0.99 | 43.70 | 0.99 | 46.22 | 3.11 |
|  | t1 | 49.09 | 2.42 | 48.75 | 2.37 | 43.70 | 1.02 | 44.01 | 1.01 | 46.39 | 3.14 |
|  | t2 | 49.47 | 2.43 | 48.74 | 2.36 | 43.70 | 1.01 | 44.33 | 1.04 | 46.56 | 3.16 |
|  | t3 | 49.84 | 2.45 | 48.75 | 2.38 | 43.70 | 1.01 | 44.65 | 1.06 | 46.73 | 3.20 |
|  | t4 | 50.22 | 2.45 | 48.75 | 2.38 | 43.69 | 1.02 | 44.97 | 1.08 | 46.91 | 3.25 |
|  | t5 | 50.59 | 2.46 | 48.75 | 2.37 | 43.71 | 1.01 | 45.29 | 1.09 | 47.08 | 3.30 |
| y | t0 | 499.84 | 26.41 | 498.26 | 26.27 | 544.15 | 18.60 | 543.39 | 17.96 | 521.41 | 31.84 |
|  | t1 | 493.55 | 26.78 | 498.25 | 26.64 | 544.18 | 18.96 | 538.82 | 18.37 | 518.70 | 32.51 |
|  | t2 | 486.99 | 27.21 | 498.11 | 26.64 | 544.09 | 18.87 | 534.23 | 18.81 | 515.85 | 33.32 |
|  | t3 | 480.45 | 27.60 | 498.05 | 26.70 | 544.14 | 18.91 | 529.67 | 19.11 | 513.08 | 34.38 |
|  | t4 | 473.73 | 28.36 | 498.24 | 26.67 | 543.96 | 19.00 | 524.99 | 19.46 | 510.23 | 35.66 |
|  | t5 | 467.08 | 28.87 | 498.13 | 26.79 | 544.16 | 18.97 | 520.64 | 20.02 | 507.50 | 37.25 |

**Table 2.** Values (mean ± SD) of variables *x and y* Directionally aligned, range normalized values (x^(n)^ and y^(n)^) at each timepoint (t0 to t5) for all four cohorts.

|  |  | Early Disease<br>Progressive |  | Early Disease<br>Stable |  | Normal<br>Stable |  | Pre_Threshold<br>Progressive |  | Total |  |
| --- | --- | --- | --- | --- | --- | --- | --- | --- | --- | --- | --- |
| Normalized<br>variable | Visit | Mean | Std.<br>Deviation | Mean | Std.<br>Deviation | Mean | Std.<br>Deviation | Mean | Std.<br>Deviation | Mean | Std.<br>Deviation |
| $x^{(n)}$ | t0 | 1.46 | 0.46 | 1.46 | 0.45 | 0.50 | 0.19 | 0.50 | 0.19 | 0.98 | 0.59 |
|  | t1 | 1.53 | 0.46 | 1.46 | 0.45 | 0.50 | 0.19 | 0.56 | 0.19 | 1.01 | 0.60 |
|  | t2 | 1.60 | 0.46 | 1.46 | 0.45 | 0.50 | 0.19 | 0.62 | 0.20 | 1.05 | 0.60 |
|  | t3 | 1.67 | 0.47 | 1.46 | 0.45 | 0.50 | 0.19 | 0.68 | 0.20 | 1.08 | 0.61 |
|  | t4 | 1.74 | 0.47 | 1.46 | 0.45 | 0.50 | 0.19 | 0.74 | 0.21 | 1.11 | 0.62 |
|  | t5 | 1.81 | 0.47 | 1.46 | 0.45 | 0.50 | 0.19 | 0.80 | 0.21 | 1.15 | 0.63 |
| $y^{(n)}$ | t0 | 0.95 | 0.26 | 0.97 | 0.26 | 0.51 | 0.18 | 0.52 | 0.18 | 0.74 | 0.32 |
|  | t1 | 1.01 | 0.27 | 0.97 | 0.26 | 0.51 | 0.19 | 0.57 | 0.18 | 0.77 | 0.32 |
|  | t2 | 1.08 | 0.27 | 0.97 | 0.26 | 0.51 | 0.19 | 0.61 | 0.19 | 0.79 | 0.33 |
|  | t3 | 1.14 | 0.27 | 0.97 | 0.26 | 0.51 | 0.19 | 0.66 | 0.19 | 0.82 | 0.34 |
|  | t4 | 1.21 | 0.28 | 0.97 | 0.26 | 0.52 | 0.19 | 0.70 | 0.19 | 0.85 | 0.35 |
|  | t5 | 1.28 | 0.29 | 0.97 | 0.27 | 0.51 | 0.19 | 0.75 | 0.20 | 0.88 | 0.37 |

A repeated-measures ANOVA on raw *x, y* values across time (*t*0– *t*5) revealed a statistically significant main effect of *time* (p < .001), and of *time* × *group* interaction for all comparison (p < .001). Plotting the raw values demonstrated diverging trajectories in progressive groups (increasing *x*, decreasing *y*) while stable and normal groups remained flat. Normalized values (*x*^(*n*)^, *y*^(*n*)^confirmed that progression occurs along a biologically plausible axis and further highlight the temporal separation between disease and stable behavior (**Figure 4)**.

**Figure 4:**
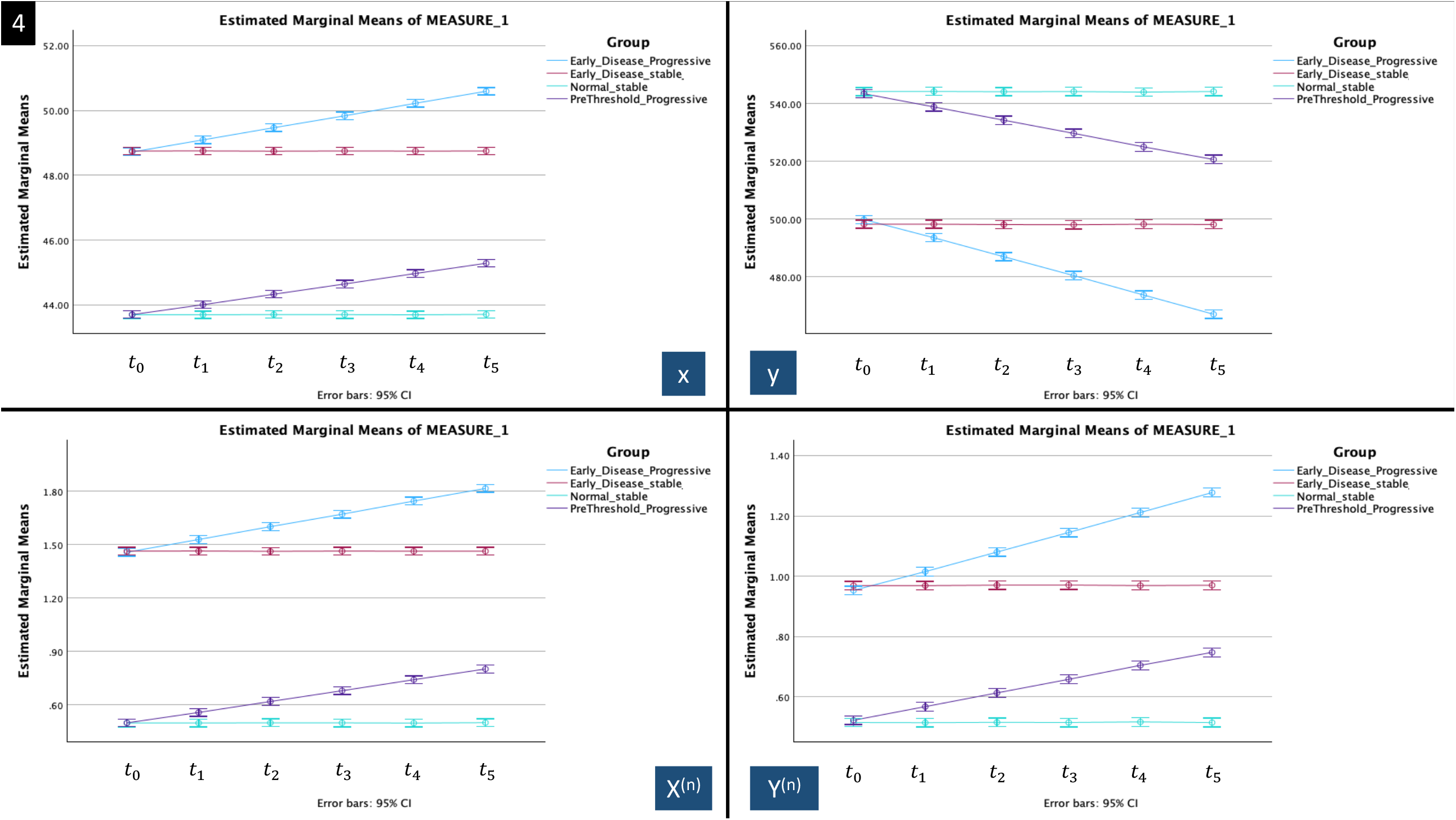
Estimated marginal means of *x* and *y* (top row) and their corresponding normalized values *x*^(*n*)^ and *y*^(*n*)^ (bottom row) across all 6 timepoints (t0-t5) for all four cohorts.

This ensured that the synthetic dataset followed the planned intention of creating 4 different datasets. The randomization and simulation did result in the trends we wanted to mimic, reassuring internal consistency.

### B: Indices’ performance

Next, we wanted to evaluate the performance of the indices we had conceptualized. The progressive groups showed a significant increase in the three metrics DEM, MNR, and CDS, whereas the stable groups did not show a significant change **(Table 3)**. Both the sets of CR derived and the Mahalanobis distance (MD) derived metrics (MNR, MNR^(MD),^ CDS, CDS^(MD)^) showed this trend. MNR^(MD)^ and CDS^(MD)^ both showed higher values for the progressives group. For the purpose of this initial study, we continue to use CR-derived MNR and CDS for the rest of the evaluation and would reserve further exploration with MD derived metrics in a higher dimensional dataset in the future.

**Table 3.** Change in of vector-based metrics DEM, MNR, and CDS from t1 to t5. Values represent mean ± SD.

|  |  | Early Disease Progressive |  | Early Disease Stable |  | Normal Stable |  | Pre_Threshold Progressive |  | Total |  |
| --- | --- | --- | --- | --- | --- | --- | --- | --- | --- | --- | --- |
|  | Follow-up | Mean | Std. Deviation | Mean | Std. Deviation | Mean | Std. Deviation | Mean | Std. Deviation | Mean | Std. Deviation |
| DEM | $t1-t0$ | 0.79 | 0.26 | 0.01 | 0.61 | -0.01 | 0.62 | 0.72 | 0.31 | 0.38 | 0.61 |
| | $t2-t0$ | 0.89 | 0.15 | 0.01 | 0.63 | 0.01 | 0.62 | 0.85 | 0.20 | 0.44 | 0.63 |
| | $t3-t0$ | 0.93 | 0.11 | 0.03 | 0.62 | 0.00 | 0.61 | 0.90 | 0.15 | 0.46 | 0.63 |
| | $t4-t0$ | 0.95 | 0.08 | 0.00 | 0.62 | 0.00 | 0.62 | 0.92 | 0.11 | 0.47 | 0.64 |
| | $t5-t0$ | 0.98 | 0.04 | 0.02 | 0.71 | 0.03 | 0.71 | 0.97 | 0.05 | 0.50 | 0.69 |
| MNR | $t1-t0$ | 0.95 | 0.38 | 0.47 | 0.25 | 0.35 | 0.18 | 0.78 | 0.33 | 0.64 | 0.38 |
| | $t2-t0$ | 1.82 | 0.55 | 0.46 | 0.24 | 0.35 | 0.18 | 1.46 | 0.51 | 1.02 | 0.75 |
| | $t3-t0$ | 2.67 | 0.67 | 0.45 | 0.25 | 0.35 | 0.18 | 2.13 | 0.62 | 1.40 | 1.13 |
| | $t4-t0$ | 3.56 | 0.79 | 0.47 | 0.25 | 0.35 | 0.19 | 2.83 | 0.73 | 1.80 | 1.52 |
| | $t5-t0$ | 4.44 | 0.91 | 0.46 | 0.25 | 0.35 | 0.18 | 3.49 | 0.82 | 2.18 | 1.92 |
| CDS | $t1-t0$ | 0.79 | 0.42 | 0.00 | 0.31 | 0.00 | 0.24 | 0.60 | 0.37 | 0.35 | 0.49 |
| | $t2-t0$ | 1.64 | 0.60 | 0.00 | 0.32 | 0.01 | 0.24 | 1.27 | 0.55 | 0.73 | 0.87 |
| | $t3-t0$ | 2.50 | 0.73 | 0.01 | 0.32 | 0.00 | 0.24 | 1.93 | 0.67 | 1.11 | 1.24 |
| | $t4-t0$ | 3.39 | 0.84 | 0.00 | 0.33 | 0.00 | 0.25 | 2.62 | 0.76 | 1.50 | 1.64 |
| | $t5-t0$ | 4.34 | 0.93 | 0.01 | 0.37 | 0.01 | 0.27 | 3.38 | 0.82 | 1.93 | 2.07 |
| DEM: Directional Emphasis Multiplier, MNR Magnitude to noise ratio, CDS Composite Drift Score |  |  |  |  |  |  |  |  |  |  |  |

As shown in **Figure 5**, the subject vector angle (top-left panel) remained stable over time, with no significant *time* × *group* interaction, reflecting that subject maintained a consistent drift direction in the *x, y* space (Repeated-measures ANOVA p>0.5). In contrast, Directional Emphasis Multiplier (DEM) increased significantly over time in progressive groups (repeated-measures ANOVA, p<0.001) (top-right panel). This may reflect improving alignment with the disease trajectory and not a large change in direction. This could also suggest the self-correcting nature of the index due to repeated measures from the baseline over follow-up. Again, as this was synthetic data, we do expect more noisiness in real cases and therefore this finding if repeated in those groups, needs to be explored further. Also, with time the pre-threshold progressive cases (PT_P) trended to match the performance of the early progressive disease (ED_P). This again is suggestive of a possible stronger alignment with confirmed disease process with time. This was not a planned or hard coded step but is explainable on disease behavior, further suggesting that even though this data was synthetic, did show patterns we expected.

**Figure 5:**
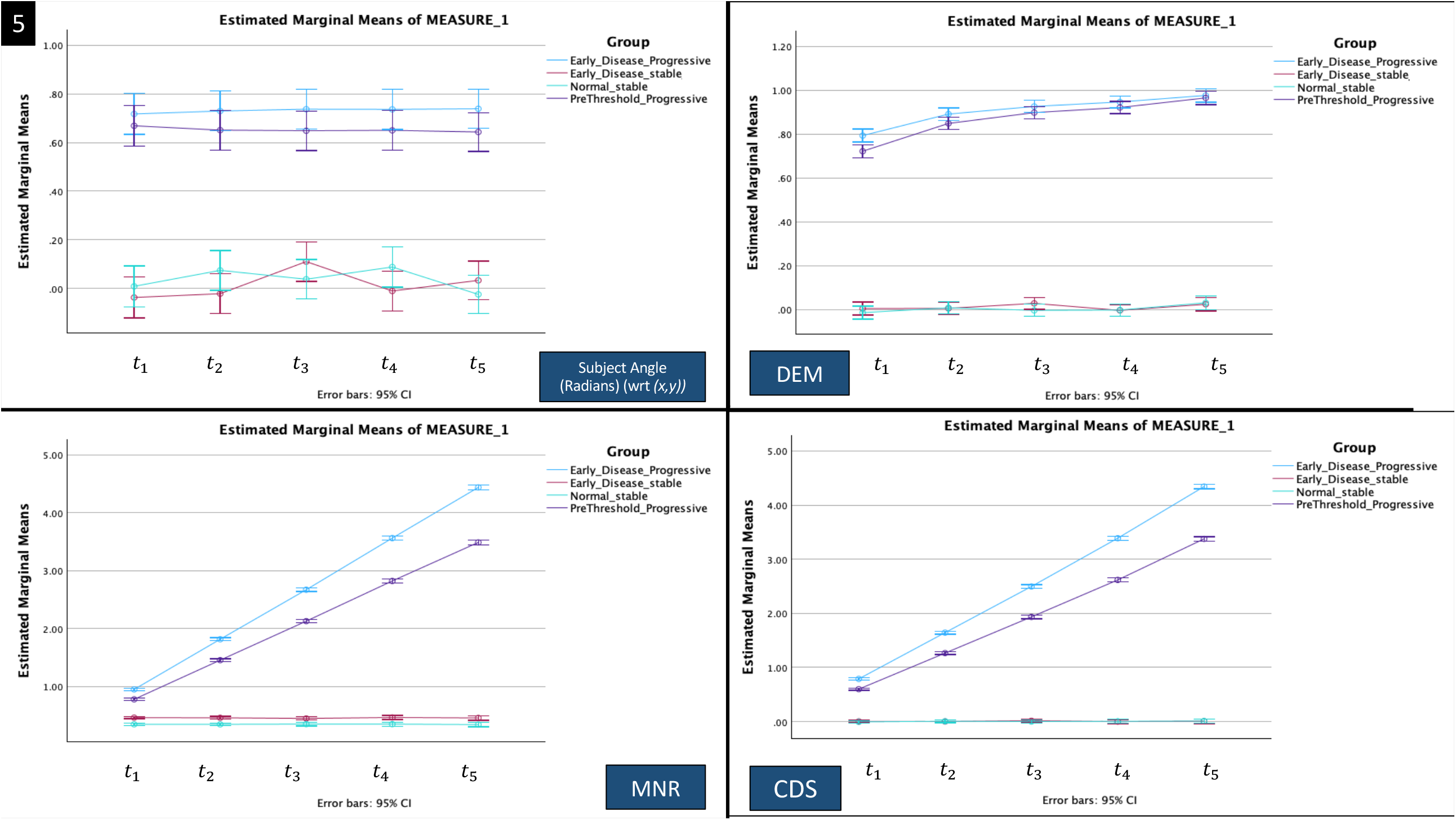
Estimated marginal means of four derived vector-based metrics over the five follow-ups (*t*1 − *t*5) grouped by cohort: Top-left: Subject vector angle (in radians) with respect to the *x, y* coordinate system. Top-right: Directional Emphasis Multiplier (DEM). Bottom-left: Magnitude-to-Noise Ratio (MNR). Bottom-right: Composite Drift Score (CDS).

The Magnitude-to-Noise Ratio (MNR) (bottom-left panel) rose in progressive groups, indicating that observed drift exceeded physiological noise boundaries (repeated-measures ANOVA, p<0.001).

Finally, the Composite Drift Score (CDS) (bottom-right panel), showed a rise in Early Disease Progressive and Pre-threshold Progressive groups, while remaining flat in stable cohorts (repeated-measures ANOVA, p<0.001).

Taken together, these vector-based metrics demonstrated strong discriminative behavior across simulated disease states, justifying the potential for further exploration for trajectory analysis and diagnostic comparison in more complicated or real-life datasets.

To visualize how directional drift evolves over time and contributes to the Composite Drift Score, we constructed a four-panel boxplot montage **(Figure 6)**. Subject angle (top-left panel) revealed that progressive groups converged directionally toward the canonical disease vector, while stable and normal groups remained without a clear directional trend. This angular trend was reflected in rising DEM values (top-right panel), which translated into increased alignment. Combined with the rising MNR (bottom-left panel), CDS values (bottom-right panel), rose in progressive cohorts ED_P and PT_P, confirming that directional alignment and supra-noise drift both contributed to the CDS scores getting higher.

**Figure 6:**
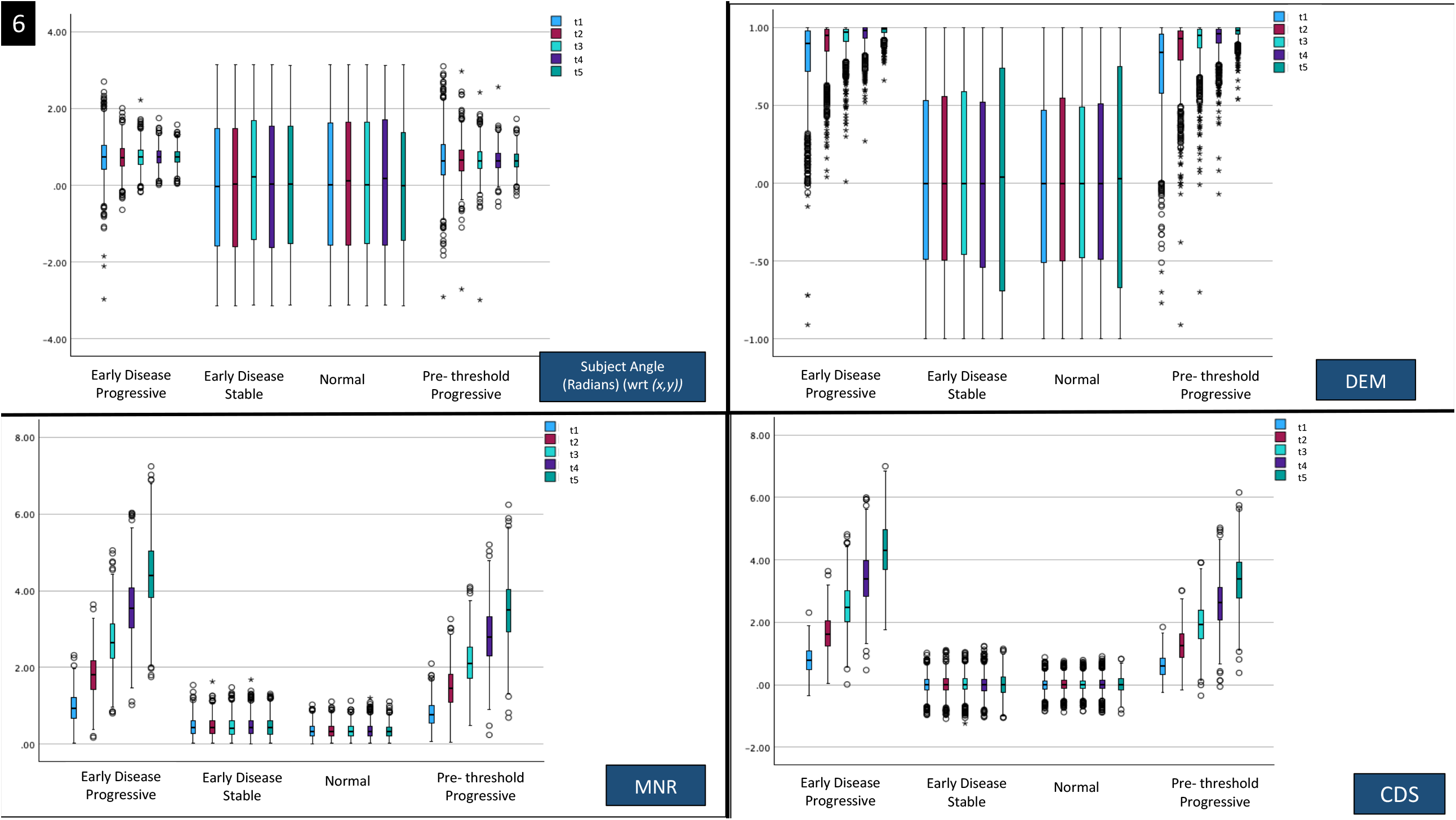
Grouped boxplots of four derived metrics over time (t1 to t5, relative to baseline t0) Top-left: Subject vector angle (in radians) with respect to the *x, y* coordinate system. Top-right: Directional Emphasis Multiplier (DEM). Bottom-left: Magnitude-to-Noise Ratio (MNR). Bottom-right: Composite Drift Score (CDS)

### C. Subject-level stochasticity visualization

Even though we induced Gaussian noise in our samples as described in the methods, at the level of measures of central tendency it is difficult to see if the outcomes created were artificially smooth without intra-individual variabilities. To assess this visually, we mapped each subject’s vector displacement over time. **Figure 7** shows directional scatter plots for all four groups across five follow-up time points, with simulated subject vectors (orange) overlaid on the baseline physiological noise cloud (blue). The stable and normal cohorts remained within the noise cloud over the follow-ups. The progressive groups showed clear but directional trends away from the noise cloud, which varied in speed between subjects. This further gave visual support to the drift varying in quantity but similar in the direction in the subjects where it was intended.

**Figure 7.**
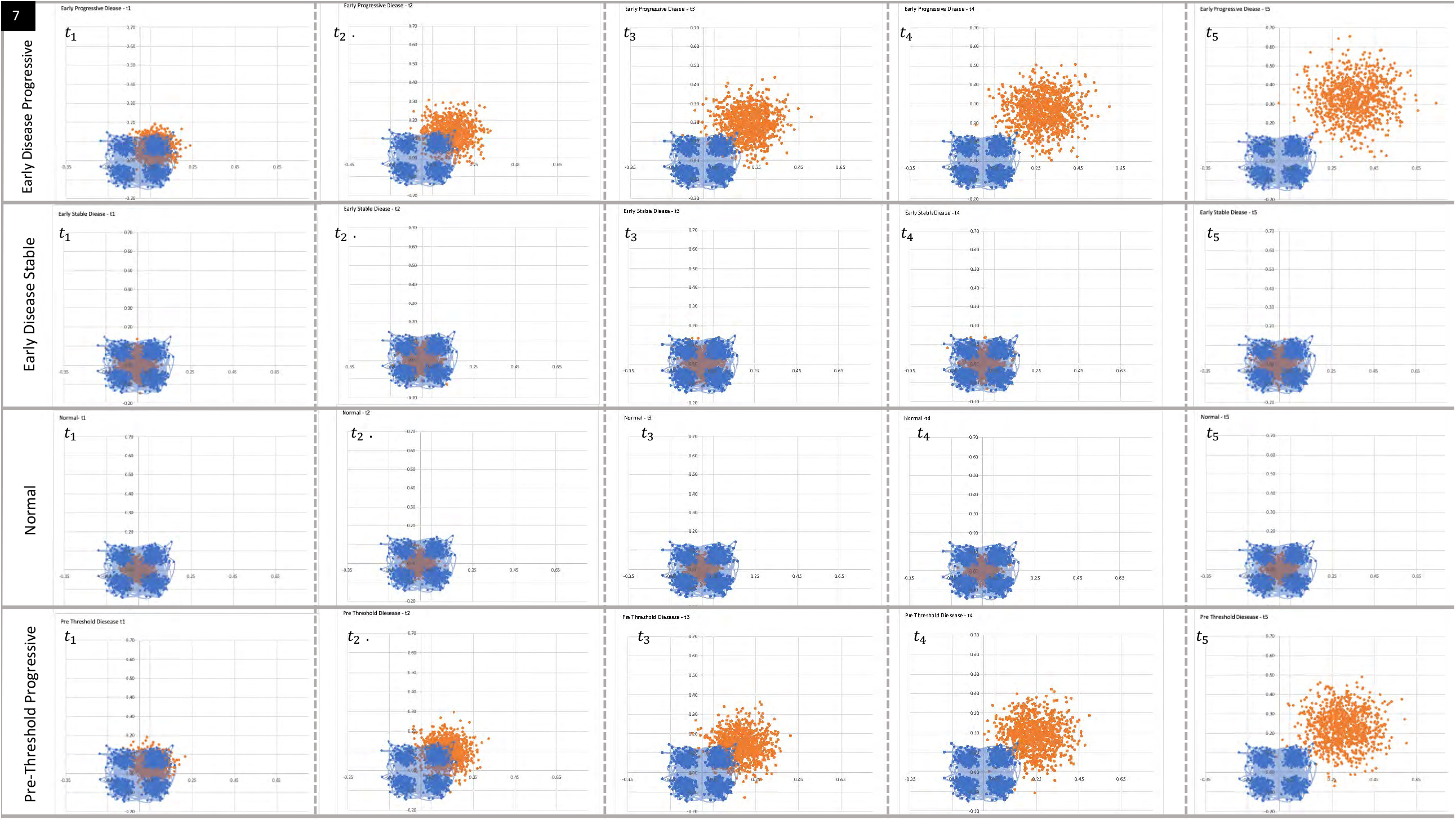
Overlay scatter plots showing subject-level displacement vectors (orange) over baseline noise cloud (blue), across cohorts and timepoints.

Perhaps, we should discuss the method of noise cloud representation in this visual analogy in more detail: As CR is a positive number, plotting the CR only would have artificially resulted in a data set limited to quadrant I of the cartesian system. In constructing the baseline physiological appearing noise cloud, we aligned the noise distribution with the biological directionality of change observed at the last visit (*i* = 5)

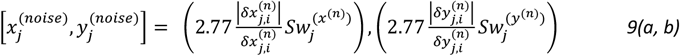

Where the 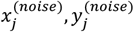are the coordinates of the noise cloud for subject 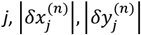 are the absolute values of 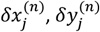, the change in the normalized *x and y* variables at the last follow-up *i* = 5, and 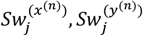are subject level intra-measurement standard deviations which when multiplied with 2.77 gives the CR for that subject. Though this approximation is not mathematically equivalent to the CR we used as a pooled value, hopefully, it gives a visual representation of the noise cloud primarily at an intuitional level. This could also be useful as we further deal with more stochastic and subject-level data in future work.

Also, **Figure 8** presents heat maps of subject-level evolution for all the follow-ups of all the cases for CDS, the cosine angle between disease and subject vectors, and magnitude of subject vector. Progressive cohorts show increasing intensity (green**→**orange**→**red) in both alignment and magnitude, consistent with disease-like drift accumulation. In contrast, stable disease and normal groups show lesser changes in the hues from the cooler colors over follow-ups. Together, these visualizations help to demonstrate the biological plausibility and internal variability of the synthetic dataset, and similar methods could be used in future work to compare the internal variability of more complicated datasets.

**Figure 8.**
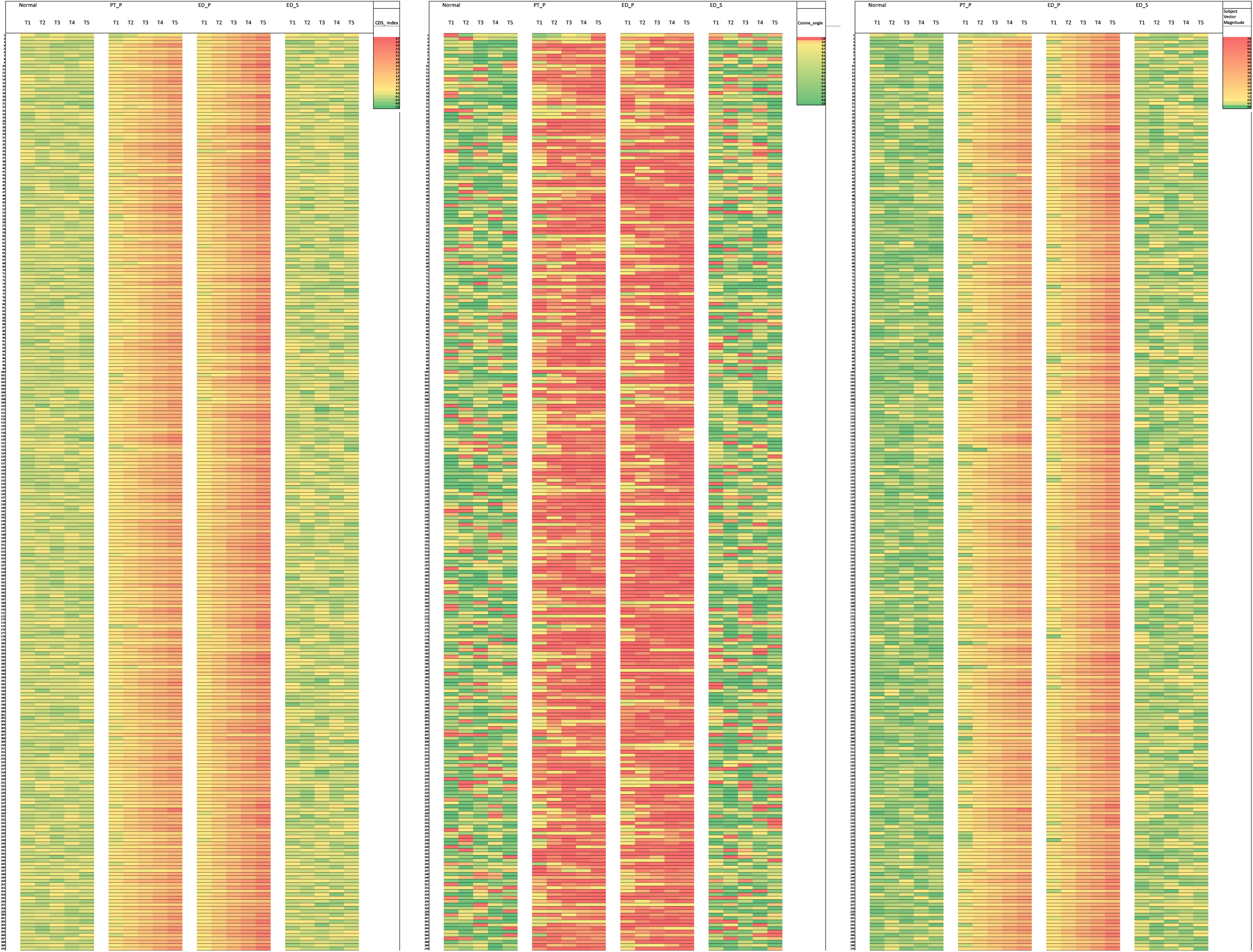
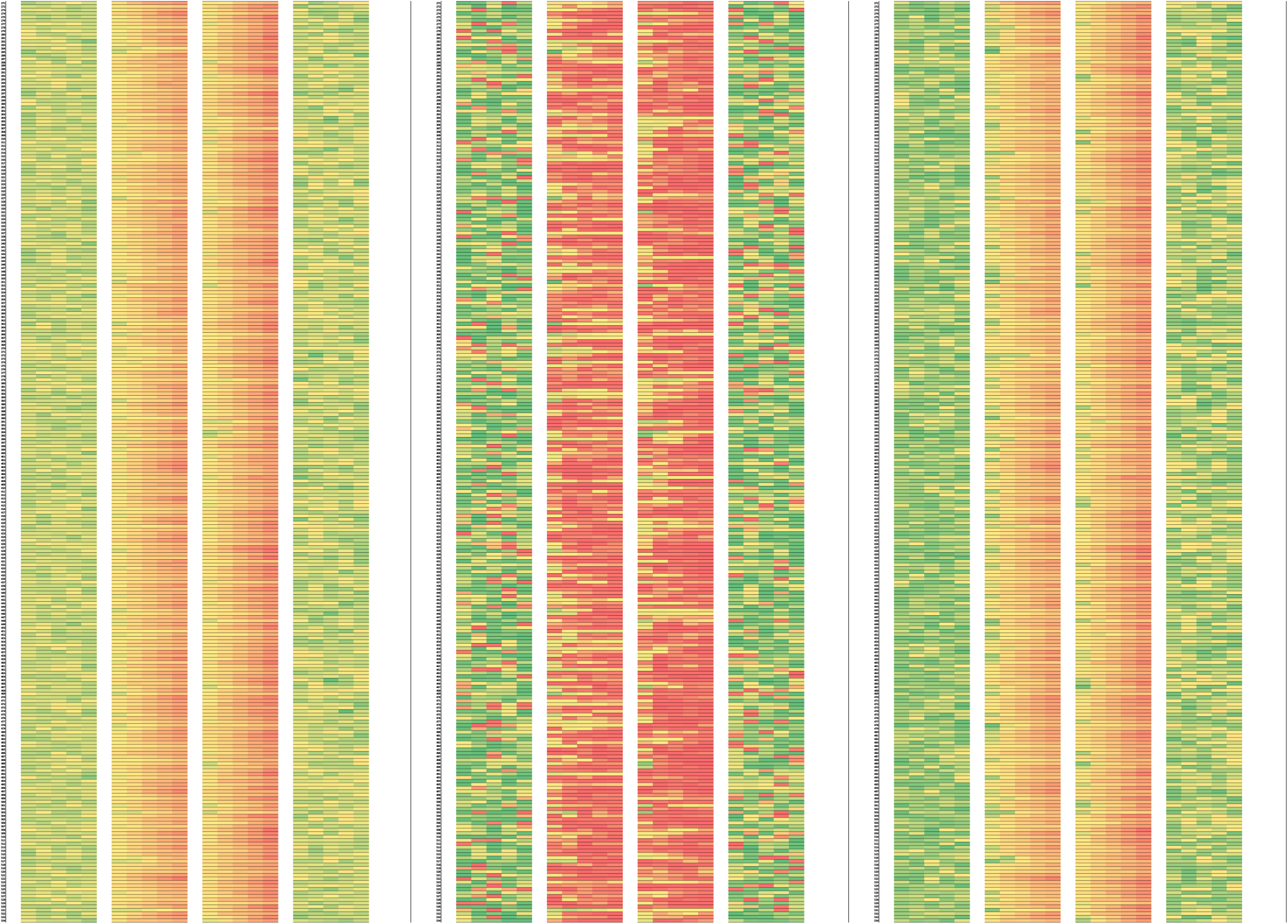
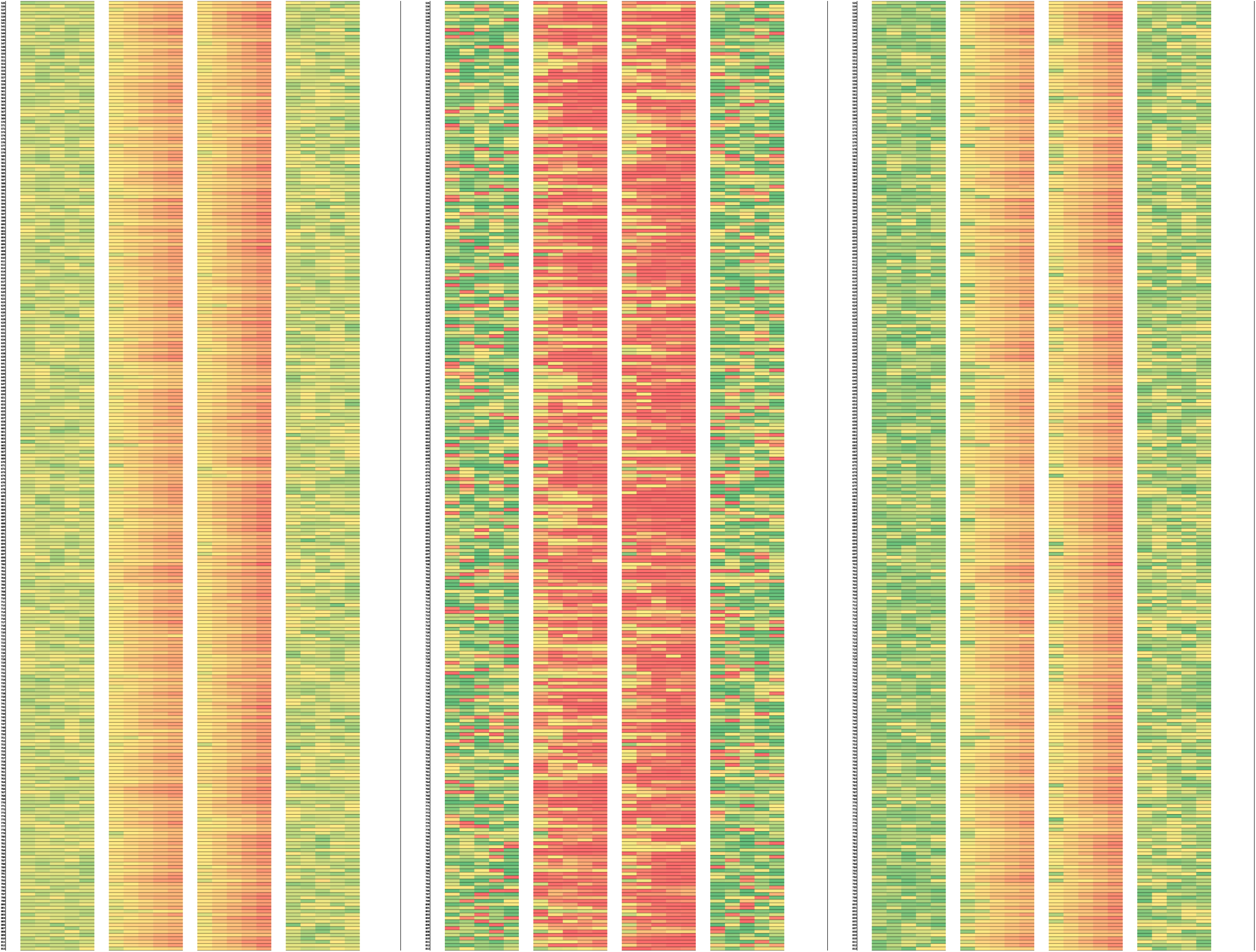
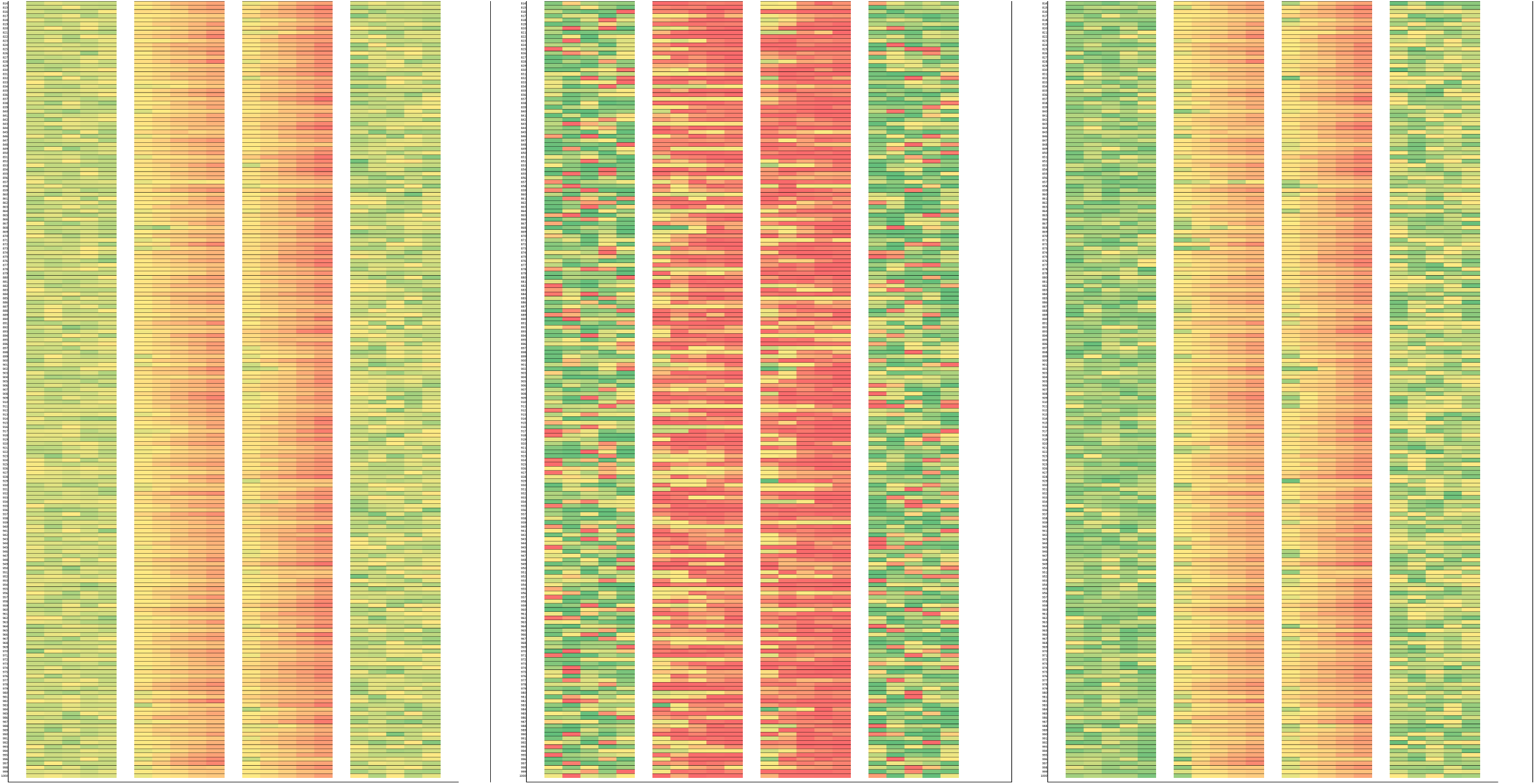
Heatmap visualization of subject-level trajectories for all the cases from each group for CDS, Cosine angle, and Subject vector magnitude across five follow-up timepoints. Rows represent individual subjects; warmer colors indicate increasing values.

## Discussion

### Background and need for this framework

This novel framework quantifies directional progression instead of relying solely on value-based thresholds. In this paper, we have conceptualized and then evaluated a geometric framework rooted in biological plausibility and clinical interpretability. The framework is derived from previous work in the fields of medicine, biomedical imaging, and signal processing. It derives inspiration from concepts in ecology, systems biology, and physics [56-59]. This is also an extension from our previous studies of univariate & bivariate range normalization, rate of change assessment and predictive modeling in keratoconus, use of repeatability indices for clinical decision-making, and modeling the role of directional alignment in the disease process [64,73-75]. The goal of this paper is to formalize a clinical intuition many physicians have when population-based cutoffs, which have historically worked well, lag in predicting patient’s unique changes [60-62].

### Attempted parsimony and interpretability

Often, correlated biomarkers lead to overfitting and interpretation difficulty [73-74]. Therefore, the parsimonious design of this model is an attempt to safeguard against overfitting. While there can be exceptions, the initial plan should be to include 2 or 3 parameters that are used for threshold-based screening. The reason to use parameters already being used commonly in threshold-based testing is to keep the clinical interpretability intact due to preexisting familiarity in clinicians for those parameters (directionality and not necessarily the numerical thresholds). Continuing from our example of keratoconus, studies have shown the advantage of corneal wavefront measurement as a more sensitive marker for keratoconus diagnosis, and Brillouin microscopy can detect keratoconus much earlier than conventional testing **[75]**. However, most ophthalmologists will know that the cornea gets thinner and steeper with keratoconus and will be able to relate to keratometry and pachymetry at a more intuitive level than corneal wavefront [4,28].

As an expansion of scope, we visualize this system to be a framework that, if proven, may be transferable to the more comprehensive practice physicians such as a primary care physician or a comprehensive ophthalmologist who can work along with subspecialists. Therefore, the common interpretability of the parameters becomes important.

The use of relatively more esoteric parameters and complex indices has an undeniable role in severity mapping for advanced diseases, but the current framework is aimed to supplement that by working in a hitherto unexplored area **[3,5,76]**.

### Concerns for oversimplification and the scope of the framework

However, one may be concerned if this is an oversimplification (using lower order, linear approximations rather than the complex black box style models). Clinicians and researchers are aware of the complex stochastic process of disease progression after the clinical thresholds are triggered up to complete loss of structure or function or both (as the case may be) [**77-79]**. Simply put, disease progression starts and stops, accelerates, and decelerates, and multiple genetic and environmental roles need to be modeled when mapping the entire disease process **[80-82]**. However, this framework does not aim to model that process but zooms in a small area of that process with an awareness of the start and end points. The starting point of this evaluation is the deviation away from the normal physiology, or as the model conceptualizes, the physiological plane. The endpoint is the crossing over of the diagnostic boundary. When the classic decision boundary is already breached, the clinician has proof of worsening, so the geometric framework does not serve any further diagnostic advantage. The patient can be flagged by either threshold-based or drift-based methods at that point in time. Linear approximation of what is possibly still a small part of the trajectory may be both mathematically acceptable and clinically translatable. **Figure 9** demonstrates this concept. The full curve is nonlinear and stochastic (blue). The dashed red line represents the actual disease trajectory within the noise-to-threshold zone, which can be locally approximated using linear assumptions. This model is not designed to fit the entire disease course but to identify early directional drift (gray arrow) away from the physiological corridor (green band). Later progression may require alternate modeling approaches, such as polynomial fitting (solid red line).

**Figure 9:**
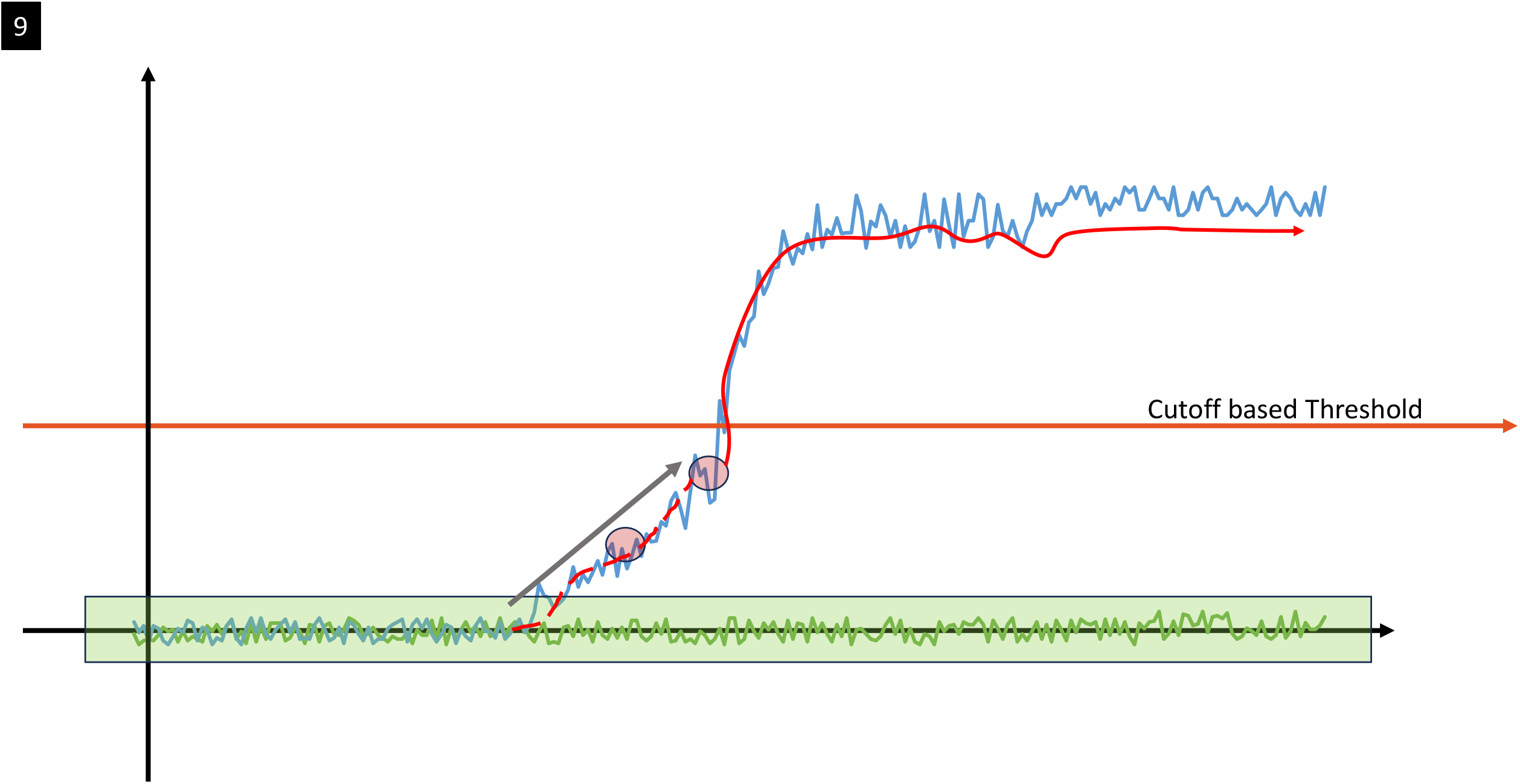
Linear approximation and considerations for the pre-threshold vs full model

For example, imagine a cornea that starts to develop keratoconus beginning at 45D (when it was structurally normal and stable) and ends at 75D (when there is an advanced, end-stage disease). The change from 45D to hitting the statistical threshold at 47.4 D is a smaller part of that curve and the endpoints can be approximated. A clinician is also interested in knowing the trend (for example, a change from 45 to 46.5 D) on a more practical basis rather than knowing if a complex polynomial can map this change. That style of fitting, as we have previously demonstrated, becomes more useful in mapping the entire disease range, from normal through early to advanced [82]. From a more methodological perspective, this idea of linear regression is derived from the practice of segmented regression analysis used to evaluate changes in interrupted time series [83].

### Deducing the start point

The visualization of the start point perhaps needs more explanation: it is the deviation away from normal in a significant, biologically plausible manner. This uses two concepts: normal noise envelope and angular alignment (cosine) with the disease process.

### Normal Noise envelope

As discussed earlier, the noise in a subject-tester-instrument-interpretation environment can be accounted for using the intra-measurement standard deviation. This extends from the classic work of Bland and Altmann [84]. This is a usual practice in many clinical domains, especially ophthalmology. Multiple previous repeatability and reliability studies have used the intra-measurement standard deviation (Sw) and its derived parameter the coefficient of repeatability (CR), especially for normal subjects [44,46,84-90]. Reference change values (RCV) used for laboratory measurements are conceptually similar [30,31]. There is an interest in using a log-normal spread for reference change value (RCV) and this model can be customized based on the set of tests it is being applied to in the future [91]. For this study, we retained the CR at 2.77 *Sw*, per the Gaussian assumption. Using the Mahalanobis distance-based metric as an alternative is very promising, especially when we would deal with ≥3-dimensional testing, or we are forced to use correlated variables. Our future studies will explore this more in Python based platforms.

When measuring multiple variables, measures of confidence can be treated as orthogonal scalars creating a vector, as has been demonstrated previously [92,93]. This is generally derived from pooled data. However, to incorporate the real-world noisiness we calculated the individual vectors for Sw between the two parameters, accounting for a creeped-in covariance, and later scaled them by 2.77 to construct what we term the “noise fog envelope”. Like the rest of our model, this is also scalable into higher dimensions.

We deliberately avoided pooling noise metrics till later steps. This method retains the option of customization for a specific subject where the noise fog envelope can be calibrated based on their noted Sw [42]. Once this envelope is modeled, it is supposed to be the upper limit of normal difference in measurement seen and therefore it was used to scale the change noted in the subject being tested. Classically, noise ellipses have been used as comparative tools (e.g., for inter and intra-device agreement). However, we have previously demonstrated their potential as a thresholding tool for clinical decision-making for cross-linking in progressive keratoconus [64]. The current metric is an extension of that concept.

### Angular alignment (cosine) with the disease process

Parameter values can change due to non-pathological reasons or due to a different pathology. For example, for a patient being evaluated for keratoconus, a tight contact lens can create a corneal warpage or scarring can cause corneal thinning. However, warpage will not induce significant corneal stromal thinning (can cause focal epithelial thickening) and corneal scarring will generally cause flattening [4,94]. So, the directionality of the pathological process becomes relevant (In our keratoconus example-corneal thinning and corneal steepening is the expected combination over time).

So, we need a method to note if the change is more than physiological noise AND is in the direction of the progression of pathology (alignment with the disease vector). This is the purpose of the combined metric, the Composite Drift Score (CDS). It combines both the magnitude and the directionality and thereby creates a potential metric to evaluate the change.

### The intuition behind the proposed initial cutoffs of CDS≥1 and for using DEM as a signed square Cosine

We propose that for initial testing, and our ongoing work, a CDS of ≥1 be considered an initial cutoff between normal and pre-threshold disease.

The best-case scenario for angular alignment can be when the angles are perfectly aligned (0 degrees between them, leading to a cosine of 1.0) and the largest-difference scenario can be diametrically opposite directions (cosine -1.0) (which may have the potential as a treatment response gauge in future iterations from this concept). As the subject’s vector drifts away from the putative disease vector, the cosine drops, penalizing a magnitude that is not directionally aligned with the disease process. Similarly, even with a perfectly aligned angle a magnitude of change smaller than noise will keep the value of the metric less than 1. As it is obvious, a large magnitude of change can overcompensate for a non-negative cosine value. For example Cos Ø =0.2 and Magnitude to noise ratio, MNR>5 would also result in the product being more than 1. To address this, we applied a transformation we called the Directional Emphasis Multiplier (DEM), instead of using the raw cosine between the subject and disease vectors. This function keeps the directional sign but gives more weight to alignment with the disease trajectory, while gradually down-weighting misaligned changes. The goal is to enhance biological interpretability by highlighting changes that are both strong and directionally consistent. For this initial study, we used the function *Cos* Ø · |*Cos* Ø|, which also be called the “signed cosine square”. It preserves the directional sign of the angle, but because the function drops off quadratically, it increasingly rewards alignment. As a simple thought experiment, at this setting of DEM, to achieve a CDS of 1, a directional misalignment of 22.5 degrees needs magnitude to be 1.2 times the noise, but this increases to 2x at 45 degrees, 5x at 67.5 degrees and infinity at 90. However, if a still tighter alignment is needed, a steeper but directionally aligned function such as (*Cos* Ø)^3^ or (*Cos* Ø. |*Cos* Ø|^W^), can be used in future datasets (**Figure 10a)**. For a constant MNR, this would result in different net CDS as seen in **Figure 10b**. CDS drops to zero at orthogonal misalignment (±90°), with all the penalty profiles depending on the chosen weighting strategy. DEM (middle row) balances directional specificity and biological smoothness, unlike the harsher Cos^3^(θ) or the more permissive Cos(θ).

**Figure 10a.**
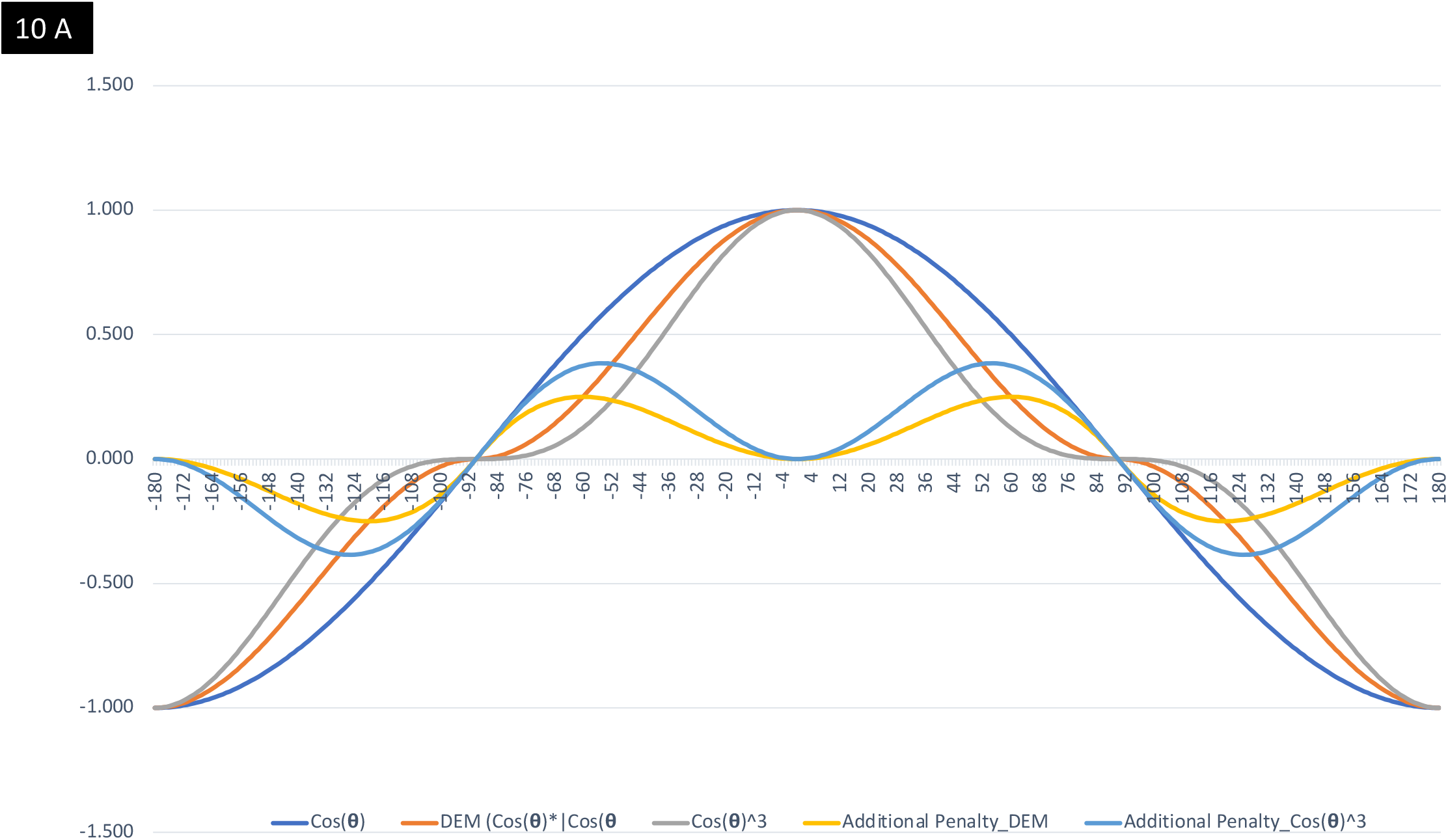
Comparison of waveforms of Angular Weighting Functions: Raw cosine (Cos(θ)), current Directional Emphasis Multiplier (DEM = Cos(θ)·|Cos(θ)|)) and a steeper cubed cosine (Cos^3^(θ)). The additional penalty values show the additional reduction each of two functions (DEM and Cubed cosine) imposes relative to Raw cosine.

**Figure 10b.**
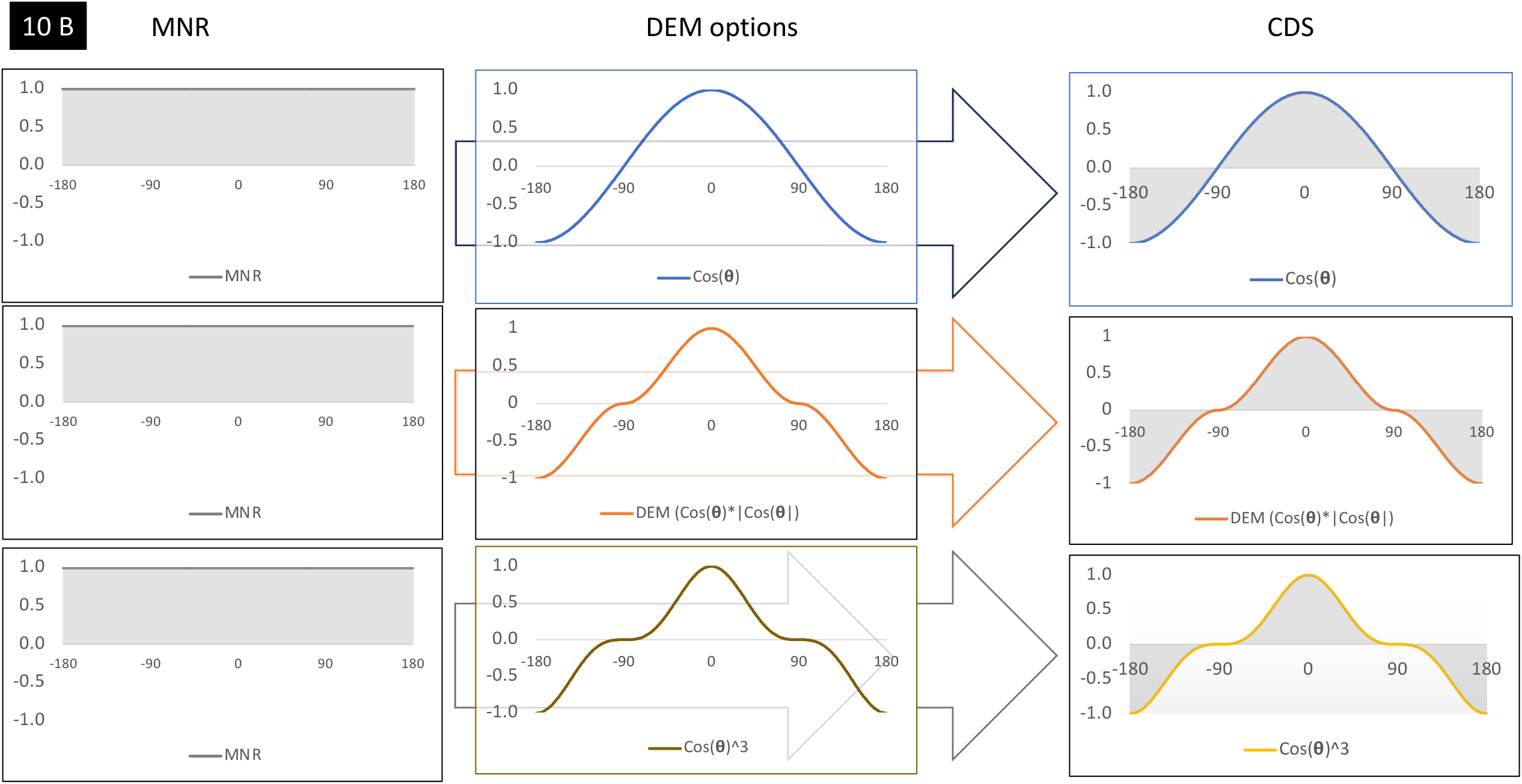
Effect of Angular Weighting on CDS Magnitude. Left: Constant magnitude-to-noise ratio (MNR). Center: Angular weighting functions: Cos(θ), DEM, and Cos^3^(θ). Right: Product of MNR and Angular function defines CDS magnitude (gray shaded area under curve).

Further optimization of directional weighting, phenotype-specific modeling, and performance tuning across noisy datasets will be addressed in subsequent work.

### Reasons for the use of Synthetic data

We should also discuss the use of synthetic data in the study. Similar approaches using synthetic or semi-synthetic disease progression models have been used previously [96-99]. There are relatively large sizes of keratoconus, Fuchs dystrophy, and normal eye databases, and we have also reported findings from our databases. However, most of these are cross-sectional [80,100-102]. Subjects who are clinically normal are often not followed up serially in a systemic fashion. Therefore, we decided to use a more customizable method such as an Excel based dataset generation vs publicly available or pre-generated cross sectional synthetic datasets in this initial explorative study.

### Conclusion

To conclude, we hypothesized a 2-plane disease vs normal model, and conceptualized a mathematical framework based on it. Then we constructed an index system, CDS (= DEM x MNR) to with the eventual target to flag early change within the subthreshold range in synthetic data. This metric and its subcomponents were able to demonstrate similar trends in progressive early disease and progressive pre-threshold groups. This suggests that with more studies, rigorous evaluation, and iterations it may be able to mathematically map its intended overarching goal: a unitless ratio based intuitive method to denote disease directional change in the pre-threshold group and therefore have impact in early detection and management of a subset of chronic diseases.

Threshold based tests are agnostic to pre-threshold movement. Therefore, the next step is to compare these metrics in unlabeled data and evaluate their performance and lead time compared to threshold-based cutoffs. This early-stage simulation study sets up ground to evaluate this metric in our next work on stress testing this on highly stochastic data, then conceptualizing case-based scenarios, leading to possible multicentric/ collaborative work in real life situations.

## Data Availability

All data produced in the present study are available upon reasonable request to the authors

